# Genetic and Phenotypic Architecture of Brain Glymphatic System

**DOI:** 10.1101/2025.03.23.25323721

**Authors:** Changhe Shi, Dongrui Ma, Shuangjie Li, Chunyan Zuo, Zhiyun Wang, Yuemeng Sun, Shasha Qi, Yuanyuan Liang, Chenwei Hao, Yanmei Feng, Zhengwei Hu, Xiaoyan Hao, Mengjie Li, Ruwei Yang, Song Tan, Chengyuan Mao, Ying Jing, Yuming Xu, Yunpeng Wang, Shilei Sun, Ole A. Andreassen

**Affiliations:** Department of Neurology, The First Affiliated Hospital of Zhengzhou University, Zhengzhou University, Zhengzhou, 450000, Henan, China; Tianjian Laboratory of Advanced Biomedical Sciences, Zhengzhou University, Zhengzhou, Henan, China; NHC Key Laboratory of Prevention and Treatment of Cerebrovascular Diseases, The First Affiliated Hospital of Zhengzhou University, Zhengzhou University, Zhengzhou, 450000, Henan, China; Henan Key Laboratory of Cerebrovascular Diseases, The First Affiliated Hospital of Zhengzhou University, Zhengzhou University, Zhengzhou, 450000, Henan, China; Sichuan Provincial Key Laboratory for Human Disease Gene Study and Rare Disease Medical Centre and Department of Neurology, Sichuan Provincial People’s Hospital, University of Electronic Science and Technology of China, Chengdu, Sichuan, China; School of Basic Medical Sciences, Zhengzhou University, Zhengzhou, Henan, 450001, China; NORMENT, Division of Mental Health and Addiction, Oslo University Hospital & Institute of Clinical Medicine, University of Oslo, Oslo, Norway; Centre for Lifespan Changes in Brain and Cognition (LCBC), Department of Psychology, University of Oslo, Oslo, Norway

**Keywords:** Brain Waste Clearance, Genetic Variants, Neuroimaging Analysis, Metabolic Health, Vascular Function, Therapeutic Targets

## Abstract

**Background:** The glymphatic system plays a crucial role in clearing metabolic waste from the brain, facilitating waste exchange between cerebrospinal fluid and interstitial fluid, and supporting brain homeostasis. However, quantifying glymphatic function has been challenging. The Diffusion Tensor Imaging Along the Perivascular Space (DTI-ALPS) method offers a non-invasive approach to assess glymphatic function by calculating an index that reflects fluid mobility within the brain. This study aimed to identify genetic variants associated with the ALPS index and explore its relationships with metabolic, immune, cognitive, and health-related phenotypes.

**Methods:** Data from 43,823 participants in the UK Biobank were analyzed. After rigorous quality control, 36,997 individuals with valid bilateral ALPS indices were included. A genome-wide association study (GWAS) was conducted to identify genetic loci linked to the ALPS index. The study also explored correlations between the ALPS index and various non-imaging traits, including cognitive performance, blood pressure, and lifestyle factors. Statistical analyses included GWAS, gene enrichment analysis, polygenic risk score validation, Cox regression, and Mendelian randomization.

**Results:** The GWAS identified 14 independent loci, encompassing 3,814 single-nucleotide polymorphisms, associated with white matter integrity, brain volume, fiber tract connectivity, inflammation, and metabolism. Key candidate genes, such as *GNA12*, *SERPIND1*, and MAPT, were linked to vascular function and neurodegenerative diseases. Enrichment analysis revealed significant roles for neuronal development, signal transduction, and metabolic pathways. The ALPS index showed significant associations with non-imaging phenotypes: higher indices correlated with better physical exercise, cognitive performance, and lower metabolic risks, while negative associations were found with smoking and excessive computer use. Polygenic risk scores confirmed these associations. Further analyses suggested that higher ALPS indices may protect against Alzheimer’s disease and multiple sclerosis.

**Conclusions:** This study represents the largest genome-wide analysis of the ALPS index to date, revealing key genetic variants that influence glymphatic function and their potential role in neurological health. The ALPS index may serve as a promising biomarker for neurodegenerative disease risk and offers new avenues for therapeutic interventions aimed at improving glymphatic clearance.

## Background

The glymphatic system is a vital pathway for the clearance of metabolic waste from the brain, operating through a combination of convective bulk flow and diffusion along concentration gradients[1]. Cerebrospinal fluid (CSF) infiltrates the brain parenchyma via perivascular spaces, facilitating the exchange of waste products with interstitial fluid[2]. This dynamic mechanism ensures efficient waste clearance but also regulates ion and neurotransmitter balance, supports nutrient transport, and modulates neuronal activity[3]. Despite the inherent challenges in quantitatively assessing glymphatic system function, the Diffusion Tensor Imaging Along the Perivascular Space (DTI-ALPS) method provides a non-invasive approach for its evaluation[4]. DTI-ALPS calculates an index reflecting brain fluid mobility by analyzing the diffusivity of water molecules along the anterior-posterior axis, particularly in projection and association fibers adjacent to the lateral ventricles, thereby serving as an indicator of glymphatic clearance capacity[5].

Current research highlights that glymphatic system function is intricately linked to various genes involved in regulating vascular permeability, CSF circulation, and the transport of metabolic waste within the brain[6]. However, large-scale genome-wide association studies (GWAS) specifically addressing the genetic basis of the glymphatic system remain unexplored. In this study, we employed an automated and unified algorithm to calculate the ALPS index in a substantial cohort of over 36,000 participants from the UK Biobank (UKBB). Through GWAS, genetic variants significantly associated with the ALPS index were identified, offering insights into their potential biological mechanisms. Alongside phenotype association analyses, the study further explored the ALPS index’s potential roles in metabolism, immunity, cognition, and other health-related traits. This research elucidates the genetic variants and phenotypes linked to glymphatic system function, underlining its possible implications in disease and providing valuable insights for future therapeutic interventions.

## Materials and methods

### Ethics

This study was conducted under approved project 221671 by the UK Biobank, following protocols sanctioned by the National Research Ethics Service Committee (http://www.ukbiobank.ac.uk/ethics/). All participants provided written informed consent.

### MRI Data Acquisition and Participant Information

Brain imaging data were collected using a Siemens Skyra 3T scanner with a 32-channel RF head coil at the UK Biobank imaging centers in Cheadle, Manchester, Newcastle, and Reading. The diffusion-weighted imaging (DWI) protocol followed standards set by the UK Biobank Imaging Working Group (http://www.ukbiobank.ac.uk/expert-working-groups). Image processing involved registration, eddy correction, and diffusion tensor imaging (DTI) tensor fitting using the Oxford FSL and mrtrix3 pipelines.[7] Data from 43,823 participants undergoing their first imaging session were analyzed.

### ALPS Index calculation

The ALPS index was calculated using a shared bash script (https://github.com/gbarisano/alps).[8] Diffusion tensor metrics (Dxx, Dyy, Dzz) were extracted from co-registered FA maps and diffusivity maps. Four 5 mm diameter spherical regions of interest (ROIs) were placed in projection and association fibers near the lateral ventricles. The ALPS index was calculated as the ratio of mean diffusivity along the x-axis to the mean diffusivity along the y- and z-axes for both hemispheres, resulting in left, right, and bilateral ALPS indices.

### Genome-Wide Association

A GWAS was performed on the mean ALPS index for both hemispheres using linear regression across 7,604,629 SNPs. Regenie (v3.4.1) [9] was used with nested ridge regression and LOCO to mitigate confounding. Quality-controlled imputed data from the Wellcome Trust Centre included variants with minor allele frequency (MAF) >1%, missingness <10%, HWE p > 1×10□¹□, and mach-r² >0.8. Covariates included age, sex, genotype array, scanner site, intracranial volume, and the top 10 genetic principal components. Phenotypes were rank-transformed, and genetic inflation and heritability were estimated using Linkage Disequilibrium SCore regression (LDSC).[10]

Post-GWAS, the LD structure, genomic risk loci, candidate single nucleotide variants (SNVs), lead SNVs, and independent significant SNVs for the ALPS index were identified using the SNV2GENE pipeline in FUMA (v1.5.2).[11] Variants were annotated for pathogenicity and regulatory effects, and cross-referenced with the GWAS Catalog to identify relevant associations.

### Conditional and Joint Association Analysis

Using Conditional and joint multiple-SNP analysis using Genome-wide Complex Trait Analysis (GCTA)-COJO,[12] conditional and joint analyses were conducted to identify independent signals within significant loci. GWAS summary statistics and LD structure from the UK Biobank reference panel were used, applying a MAF threshold of 1% and a 10 Mb LD window. Independent SNPs were identified with an r² < 0.6, and lead SNPs with r² < 0.1.

### Candidate Gene Identification and Annotation

FUMA (v1.5.2)[11] was employed for positional mapping, gene-based association using Multi-marker Analysis of GenoMic Annotation (MAGMA),[13] and variant annotation with ANNOVAR.[14] Genomic risk loci were defined based on independent signals and proximity. Candidate genes were cross-referenced with the GWAS Catalog and annotated for pathogenicity and regulatory effects. Genes supported by multiple lines of evidence were prioritized.

### Transcriptome-Wide Association

We employed FUSION software (http://gusevlab.org/projects/fusion/) to construct a gene expression prediction model based on GWAS data[15]. This approach integrated gene expression data with reference panel weights for conducting a transcriptome-wide association study (TWAS). We analyzed expression weights derived from the Genotype-Tissue Expression project version 8 (GTEx v8) across 22 tissues relevant to the glymphatic system, including the central nervous system, blood vessels, blood, and peripheral nerve tissues. Using LD reference data from the 1000 Genomes Project Phase 3, we assessed the association between gene expression and the ALPS Index. To address the issue of multiple testing, Bonferroni correction (P < 0.05 / [number of genes * number of tissues]) was applied to identify significantly associated genes and their expression quantitative trait loci (eQTLs).

### Colocalization Analysis

COLOC (v5.1.0)[16] was utilized within FUSION to evaluate the co-localization of GWAS SNPs with eQTL/ splicing quantitative trait loci (sQTL) signals in a 1.5 Mb window around significant loci. A posterior probability (PP4) >80% indicated co-localization, suggesting shared causal variants.

### Functional Annotation of Susceptible Genes

We further sought to identify candidate genes influencing ALPS Index phenotypic variation using an integrative approach supported by multiple lines of evidence. Genes annotated by more than three methods were classified as candidate genes for ALPS Index. Gene enrichment analysis was performed using DAVID (https://david.ncifcrf.gov/),[17] focusing on specific Gene Ontology (GO) categories with a false discovery rate (FDR) < 0.05 after Benjamini-Hochberg correction.

### Cell-Specific Susceptibility Analysis

The scDRS algorithm[18] linked single-cell RNA sequencing data to polygenic risk for the ALPS index. Gene sets associated with the ALPS index were constructed, and susceptibility scores were calculated and normalized for each cell, with p-values derived from comparison to control scores.

### Disease Association Analysis

Enrichr (https://maayanlab.cloud/Enrichr/)[19] was used to assess associations between candidate genes and known diseases using the GWAS Catalog 2023. Enrichment significance was determined with FDR < 0.05.

### Enrichment of Drug Target Genes

We sourced druggable genes mapped to Entrez IDs from the DGIdb database,[20] extracting 5,012 potential targets (Supplementary Table 1). Additionally, we utilized Finan et al.,[21] identifying 4,463 druggable genes (Supplementary Table 2) linked to GWAS loci of complex diseases. To ensure reliability, we further screened 2,587 genes validated by both sources and assigned official names by the Human Genome Nomenclature Committee (HGNC). DrugBank[22] and ClinicalTrials (https://www.clinicaltrials.gov) databases were consulted to evaluate drug development status.[22]

### Association Analysis Between ALPS Index and Five Major Categories of Non-Imaging Phenotypes

We used the ALPS index, rank-transformed from 36,997 participants, to perform phenotype association analysis with 2,121 UK Biobank risk factors across five categories: Biomarkers, Medical Conditions, Cognitive Health, Lifestyle, and Sex-Specific Factors. Variables were sourced from Additional Exposures, Assessment Centre, Biological Samples, Health-Related Outcomes, and Online Follow-Up.

PheWAS was conducted using the PHESANT R package,[23] which classified variables as continuous, ordered/unordered categorical, or binary. In the “ALPS Index-Association Analysis” step, the rank-transformed ALPS index was the independent variable, with factors as dependent variables, adjusting for age, sex, and assessment center. We reported standardized regression coefficients (β) for continuous outcomes and log odds ratios (OR) for binary outcomes. All analyses were two-sided and corrected for multiple testing using FDR. Associations with pFDR < 0.05 were selected for further validation.

### Polygenic Risk Score Validation of ALPS Index Associations Across Five Phenotype Categories

In the validation process, we calculated the polygenic risk score (PRS) for the ALPS index in 370,920 UKBB participants who did not have a directly measured ALPS index. After excluding SNPs with a call rate below 95% and MAF below 0.1%, and selecting individuals of recent British ancestry without close relatives, we used PRS-CS[24] with GWAS data from 36,111 White British individuals to derive the PRS.

In the “ALPS Index-Association Analysis” step, the ALPS index-PRS was the independent variable, and factors with pFDR < 0.05 were the dependent variables. We adjusted for age, sex, genotyping array, the top 10 genetic principal components, and assessment center. This “ALPS Index-PRS-PheWAS” was conducted using the PHESANT package in R, employing standardized β for linear models and log-transformed OR for binary outcomes. All 181 association results were subjected to FDR correction.

### Validation with Neurological Diseases Using Mendelian Randomization and Cox

To validate the significant negative associations, we used multivariable Cox proportional hazards models to assess the relationship between the ALPS Index and the incidence of neurological diseases (Alzheimer’s disease [AD], Multiple sclerosis [MS], cerebral infarction, stroke). Follow-up started at the second recruitment visit and continued until diagnosis, death, loss to follow-up, or last hospital admission. Models were adjusted for age, sex, Townsend deprivation index, and BMI, with the proportional hazards assumption tested using Schoenfeld residuals. Participants with prior neurological diseases were excluded. Additionally, two-sample Mendelian Randomization (MR)[25] was conducted using inverse-variance weighted (IVW) and other robust methods to explore causal relationships between the ALPS Index and neurological diseases, employing independent genetic instruments and performing sensitivity analyses for heterogeneity and pleiotropy. Detailed methods are provided in Supplementary Methods.

## Results

### ALPS Index in 36,997 UK Biobank Participants

Fig. 1 illustrates the overarching framework of our study. Among the 43,823 UKBB participants with post-processed DTI data, the average diffusivity along the x-axis in the projection and association fibers (Dxproj and Dxassoc), as well as the average diffusivity along the y-axis (Dyproj) and z-axis (Dzassoc), were extracted. This allowed for the computation of the left, right, and bilateral mean ALPS indices (see Supplementary Fig. 1 and Methods section). Following stringent quality control and genetic analyses, a total of 36,997 UKBB participants with bilateral mean ALPS indices were retained for further examination. To achieve normal distribution of the ALPS index, a rank-based inverse normal transformation was applied prior to analysis.

**Figure 1.**
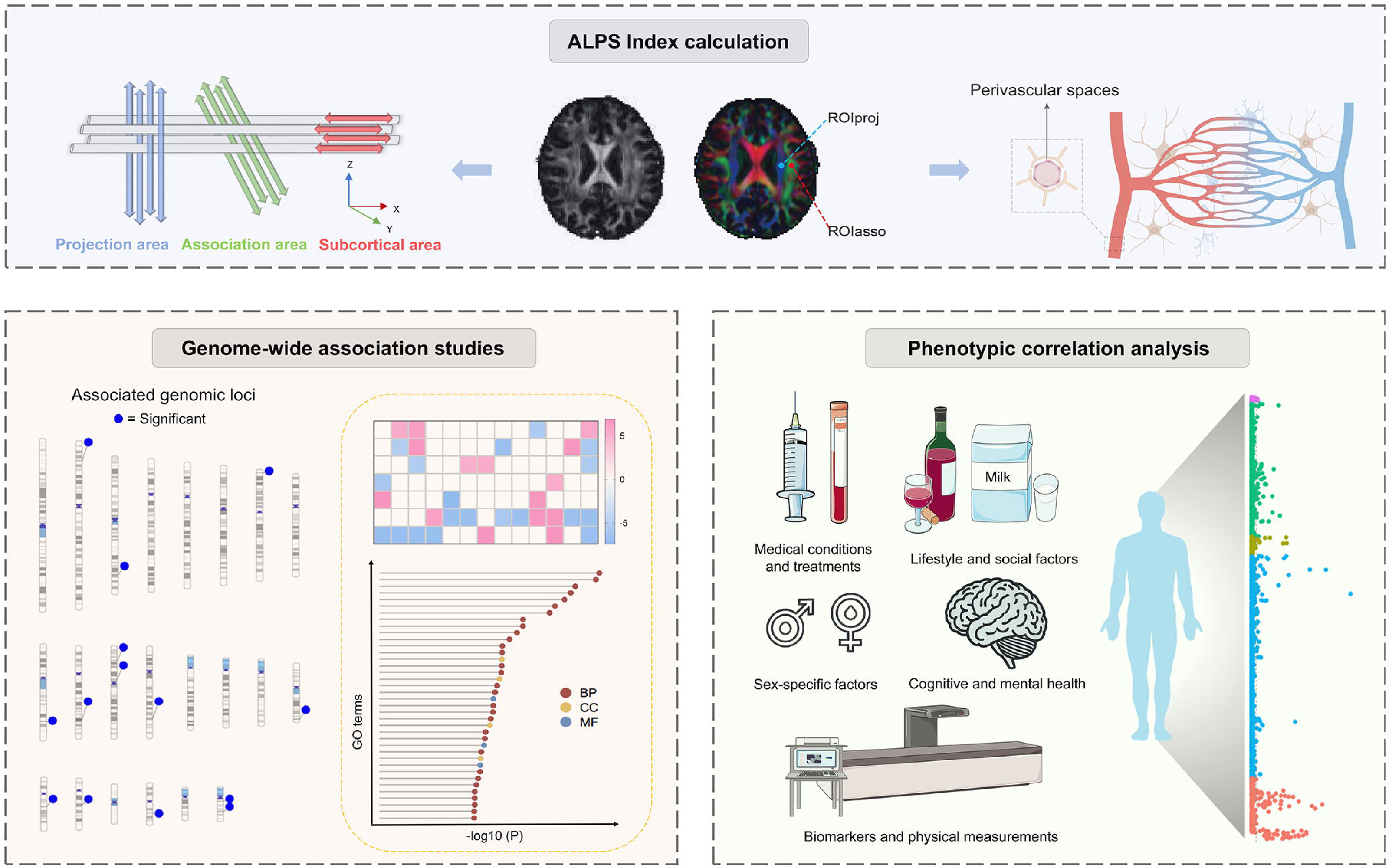
Study overview. The top section outlines the process of calculating the ALPS Index. The bottom-left section presents the GWAS analysis and subsequent analyses such as transcriptome-wide association studies (TWAS) and biological function analysis of the identified genes. The bottom-right section focuses on exploring the phenome-wide associations of the identified genes. Ideogram of associated genomic loci was generated using PhenoGram (http://visualization.ritchielab.org/phenograms/plot).

### Variants Associated with ALPS index

To ensure the integrity of our cohort, participants with neurological or psychiatric disorders (ICD-10 codes F or G) were excluded from the analysis. Consequently, the study encompassed ALPS index data from 36,111 white British participants for GWAS analysis. The mean age of this sample was 64 years (SD: 7.69; range: 45–83 years), with females constituting 52.73% of the cohort.

Using the FUMA platform, 14 independent genome-wide significant loci were identified (Table 1), associated with white matter integrity, fiber tract connectivity, brain volume, inflammation, and metabolism (Supplementary Table 3). These loci encompassed 3,814 significant SNPs linked to the ALPS index (Fig. 2, Table 1, Supplementary Fig. 2, and Supplementary Table 4). No genomic inflation was detected (Supplementary Fig. 3). Most significant SNPs were intronic or intergenic (Supplementary Table 4), with only 51 exonic variants. Additionally, 108 SNPs had deleterious Combined Annotation Dependent Depletion (CADD) scores (> 12.37) and 63 were in open chromatin regions (commonChrState of 1 or 2). LDSC analysis showed a genomic inflation factor (λ_GC_) of 1.16 and heritability of 0.26. The LDSC intercept was 1.01 with an attenuation ratio of 0.07, indicating inflation was due to polygenicity rather than population stratification, ensuring robust findings.

**Figure 2.**
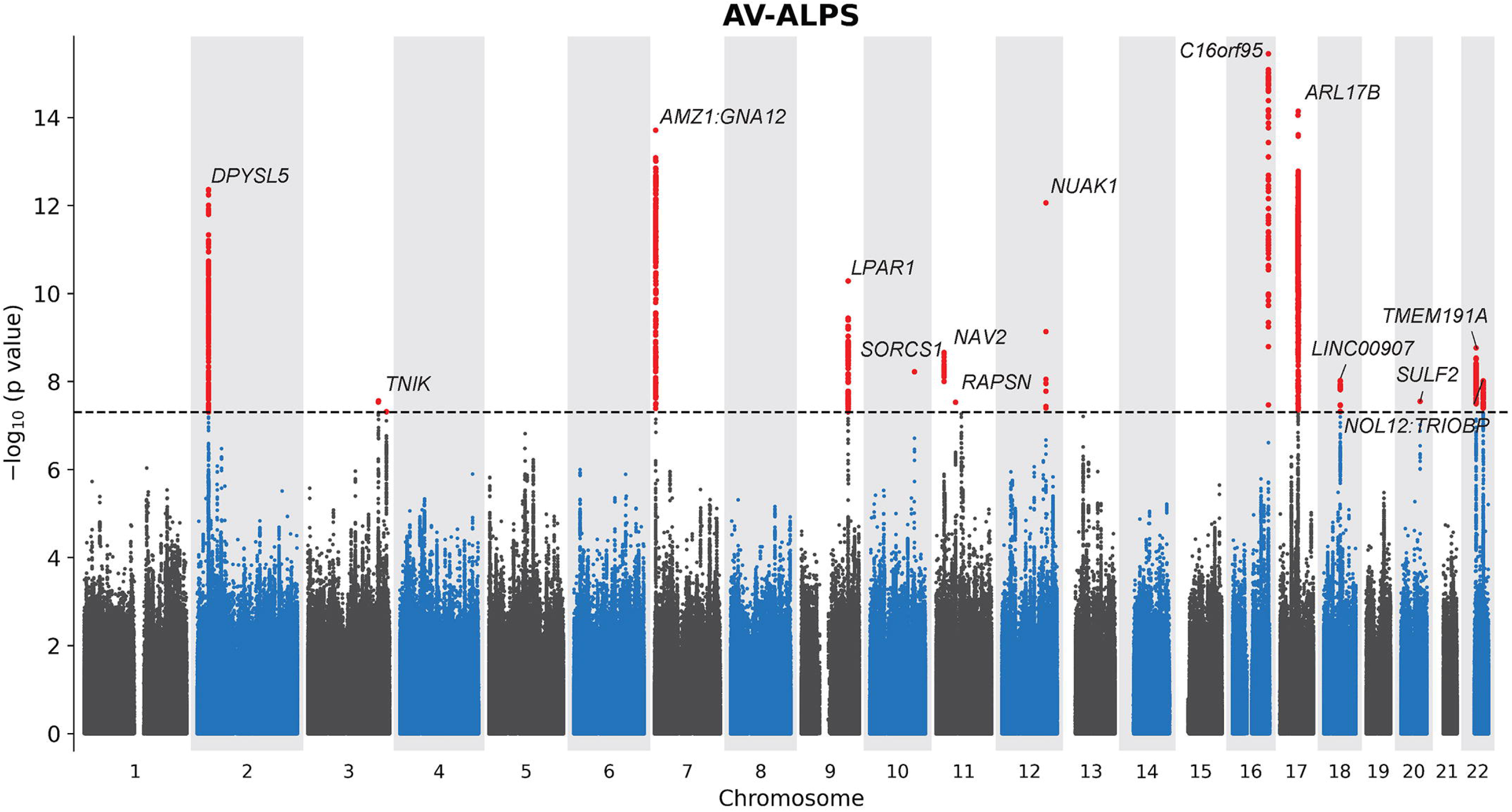
Manhattan plot showing the GWAS results. Manhattan plots showing the GWAS results derived from a two-step linear mixed model using the regenie software, with ridge regression in step one and linear regression in step two. The horizontal axis indicates chromosomal position. The vertical axis indicates −log10(P value) of the association. The dotted lines indicate the genome-wide-significance threshold of P=□5□×□10^−8^. Gene, associated gene in the identified genomic locus, the nearest Gene of the SNP based on ANNOVAR annotations. AV-ALPS, average of the left and right ALPS index. P-values are two-sided and unadjusted for multiple testing.

**Table 1.**
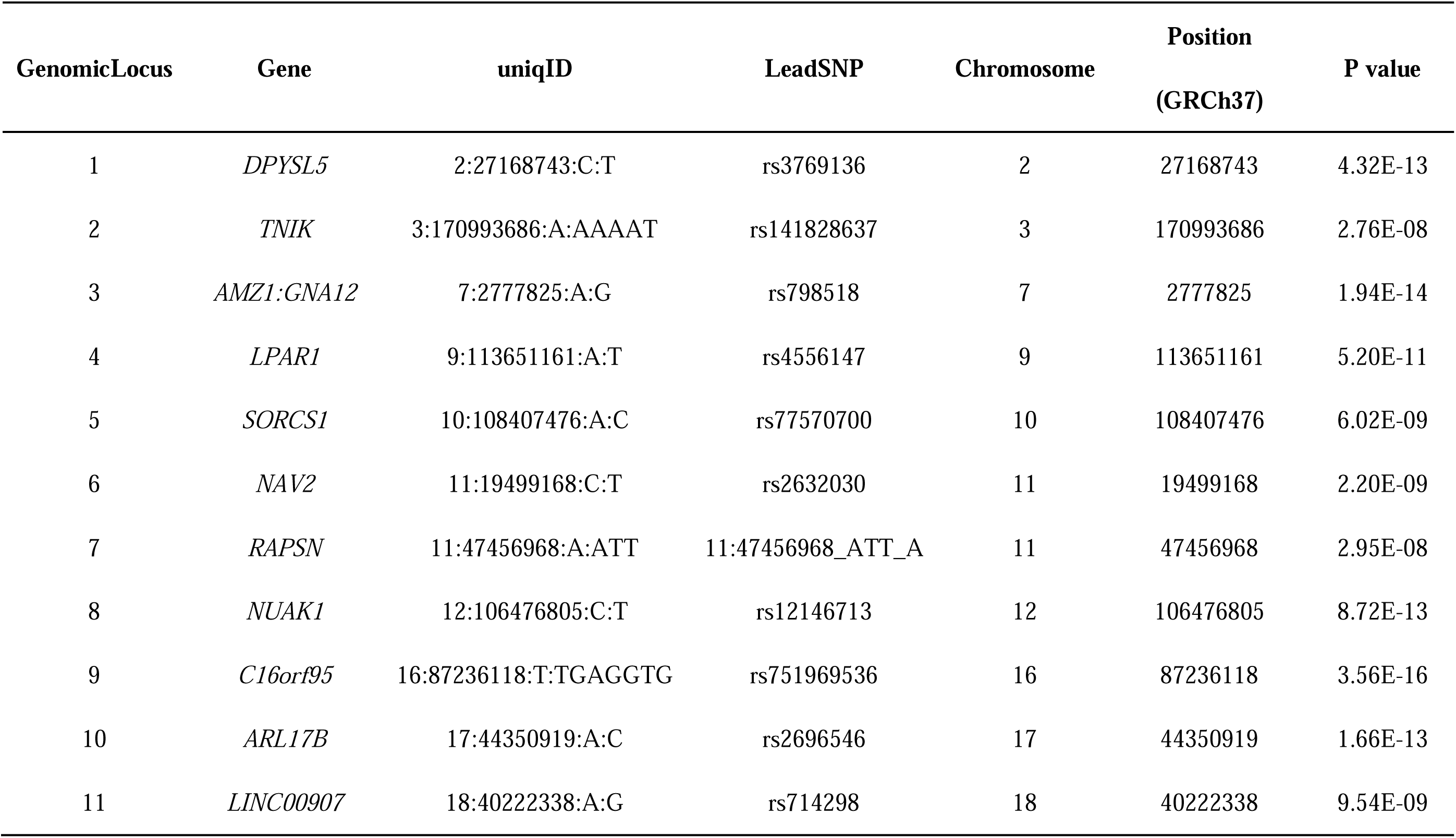
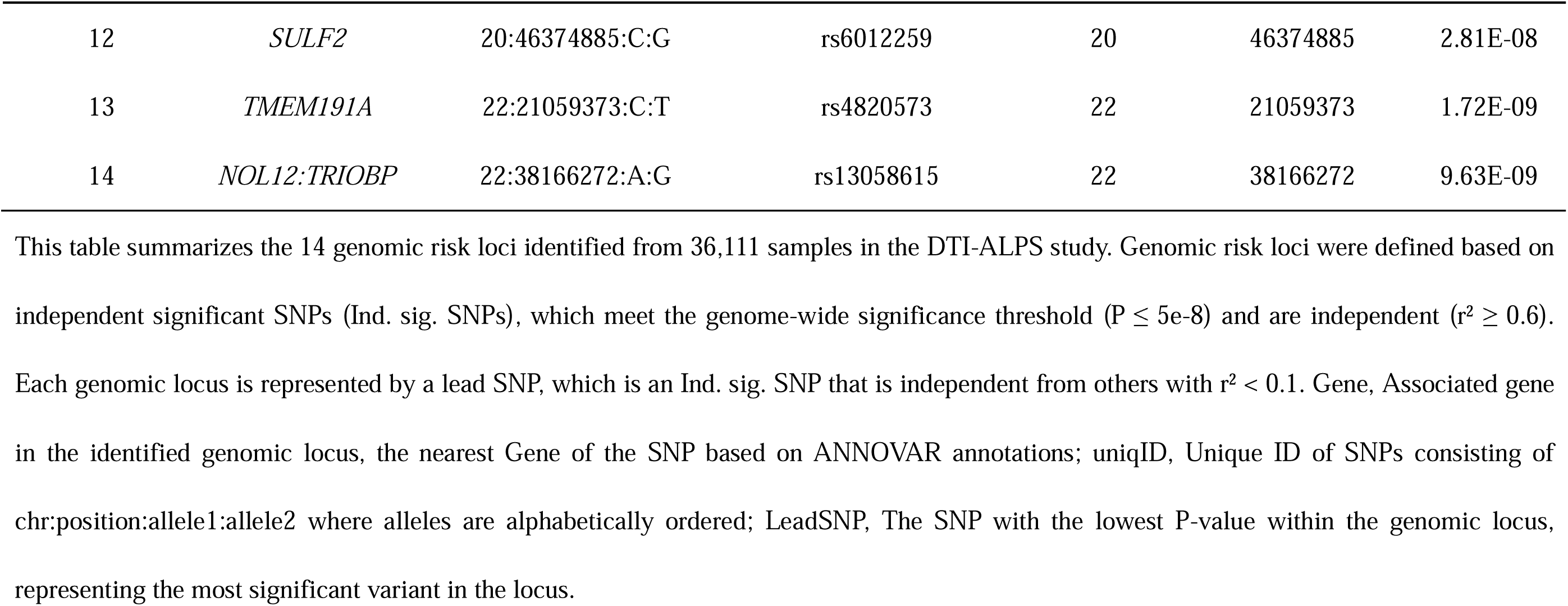
14 genomic risk loci identified with ALPS Index from 36,111 samples.

| GenomicLocus | Gene | uniqID | LeadSNP | Chromosome | Position<br>(GRCh37) | P value |
| --- | --- | --- | --- | --- | --- | --- |
| 1 | *DPYSL5* | 2:27168743:C:T | rs3769136 | 2 | 27168743 | 4.32E-13 |
| 2 | *TNIK* | 3:170993686:A:AAAAT | rs141828637 | 3 | 170993686 | 2.76E-08 |
| 3 | *AMZ1:GNA12* | 7:2777825:A:G | rs798518 | 7 | 2777825 | 1.94E-14 |
| 4 | *LPAR1* | 9:113651161:A:T | rs4556147 | 9 | 113651161 | 5.20E-11 |
| 5 | *SORCS1* | 10:108407476:A:C | rs77570700 | 10 | 108407476 | 6.02E-09 |
| 6 | *NAV2* | 11:19499168:C:T | rs2632030 | 11 | 19499168 | 2.20E-09 |
| 7 | *RAPSN* | 11:47456968:A:ATT | 11:47456968_ATT_A | 11 | 47456968 | 2.95E-08 |
| 8 | *NUAK1* | 12:106476805:C:T | rs12146713 | 12 | 106476805 | 8.72E-13 |
| 9 | *C16orf95* | 16:87236118:T:TGAGGTG | rs751969536 | 16 | 87236118 | 3.56E-16 |
| 10 | *ARL17B* | 17:44350919:A:C | rs2696546 | 17 | 44350919 | 1.66E-13 |
| 11 | *LINC00907* | 18:40222338:A:G | rs714298 | 18 | 40222338 | 9.54E-09 |
| 12 | *SULF2* | 20:46374885:C:G | rs6012259 | 20 | 46374885 | 2.81E-08 |
| 13 | *TMEM191A* | 22:21059373:C:T | rs4820573 | 22 | 21059373 | 1.72E-09 |
| 14 | *NOL12:TRIOBP* | 22:38166272:A:G | rs13058615 | 22 | 38166272 | 9.63E-09 |

This table summarizes the 14 genomic risk loci identified from 36,111 samples in the DTI-ALPS study. Genomic risk loci were defined based on independent significant SNPs (Ind. sig. SNPs), which meet the genome-wide significance threshold ( $P \leq 5e-8$ ) and are independent ( $r^2 \geq 0.6$ ). Each genomic locus is represented by a lead SNP, which is an Ind. sig. SNP that is independent from others with $r^2 < 0.1$ . Gene, Associated gene in the identified genomic locus, the nearest Gene of the SNP based on ANNOVAR annotations; uniqID, Unique ID of SNPs consisting of chr:position:allele1:allele2 where alleles are alphabetically ordered; LeadSNP, The SNP with the lowest P-value within the genomic locus, representing the most significant variant in the locus.

GCTA-COJO identified 15 independent genome-wide significant SNPs (MAF□≥□0.01, P□<□5×10□□). Fourteen of these SNPs were within the FUMA-identified loci, and one was outside (Supplementary Tables 3 and 5).

### Identification and Functional Exploration of Candidate Genes Related to ALPS Index

By analyzing GWAS summary statistics, we identified genes using four methods: positional mapping identified 46 genes within ±10 kb of lead variants (Supplementary Table 6), MAGMA gene analysis revealed 45 significant genes (mean χ² statistic, P < 0.05/19141 = 2.61 × 10□□) (Supplementary Table 7), TWAS uncovered 56 genes (Fig. 3 and Supplementary Table 8), and Bayesian colocalization integrated eQTL and sQTL data to identify 143 genes (Fig. 3 and Supplementary Table 9). Combining these methods yielded 206 candidate genes, with 11 genes consistently identified by all four methods, 14 by three methods, and 48 by two methods (Fig. 4a and Supplementary Table 10).

**Figure 3.**
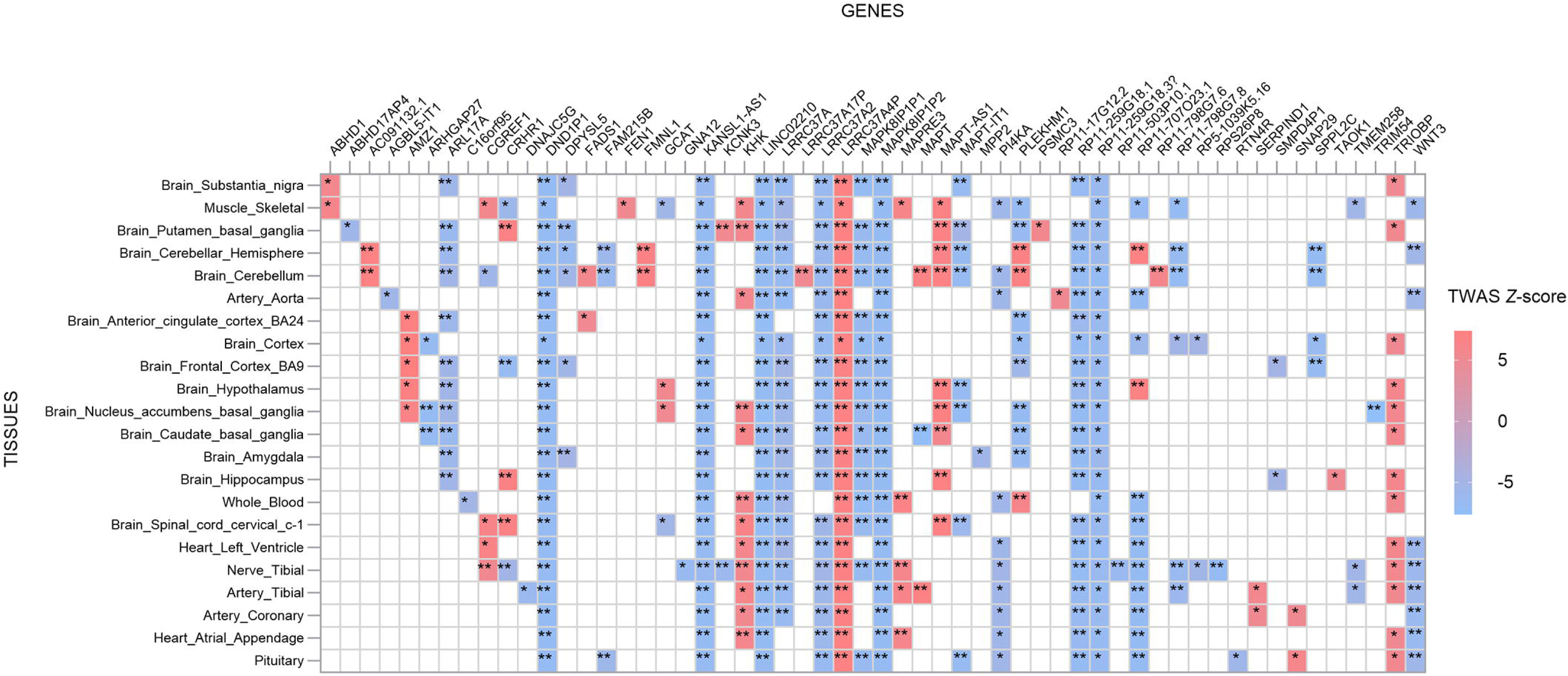
Transcriptome-wide significant genes with ALPS Index. We used precomputed functional weights from 22 publicly available gene expression reference panels from GTEx v8. Transcriptome-wide significant genes and the corresponding eQTLs were determined using Bonferroni correction based on the number of features tested in different tissues: Artery_Aorta (7,817), Artery_Coronary (3,898), Artery_Tibial (9,561), Brain_Amygdala (2,604), Brain_Anterior_cingulate_cortex_BA24 (3,451), Brain_Caudate_basal_ganglia (5,036), Brain_Cerebellar_Hemisphere (6,091), Brain_Cerebellum (7,271), Brain_Cortex (5,608), Brain_Frontal_Cortex_BA9 (4,517), Brain_Hippocampus (3,547), Brain_Hypothalamus (3,543), Brain_Nucleus_accumbens_basal_ganglia (4,988), Brain_Putamen_basal_ganglia (4,283), Brain_Spinal_cord_cervical_c-1 (3,112), Brain_Substantia_nigra (2,257), Heart_Atrial_Appendage (6,740), Heart_Left_Ventricle (5,886), Muscle_Skeletal (8,515), Nerve_Tibial (11,274), Pituitary (6,177), Whole_Blood (7,980). *Significant result in the TWAS. **significant result in the TWAS and conditional analyses, and with a COLOC PP4□>□0.8.

**Figure 4.**
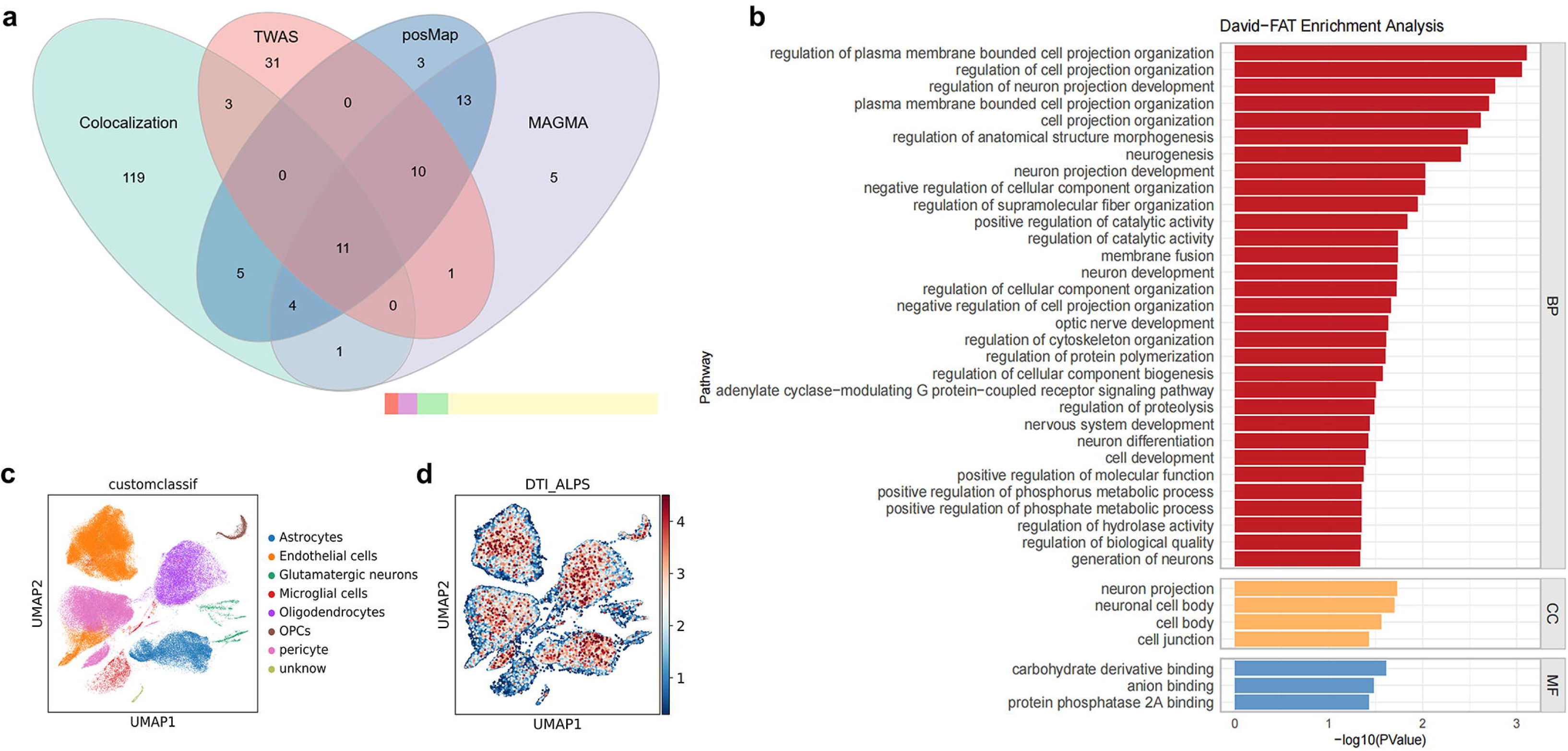
Post-GWAS analysis results. a: Annotation of candidate genes by using four different approaches. The Venn diagram illustrates the overlap of candidate genes for the ALPS index identified by four methods (Colocalization, TWAS, posMap, MAGMA). The bar chart in the lower left corner represents the proportion of annotated genes at the end of systole using these four methods: red indicates genes annotated by all four methods, purple indicates genes annotated by three methods, green indicates genes annotated by two methods, and yellow indicates genes annotated by only one method. TWAS, transcriptome-wide association study; MAGMA, multi-marker genome-wide annotation analysis. b: Gene set enrichment analysis of significant genes identified in the GWAS, using Gene Ontology and KEGG Ontology database. The p-values reported are two-sided and unadjusted. c: UMAP visualization displays clusters of human brain vascular cells, labeled according to cell types annotated with marker genes from previously published studies. d: Subpopulations of human brain vascular cells associated with ALPS Index. Cells with significant associations are marked in red, representing scDRS disease scores; other cells are shown in blue.

Several of these genes are closely linked to vascular function and CSF flow. For instance, *GNA12* regulates smooth muscle contraction and supports endothelial integrity[26], while *SERPIND1* is involved in blood coagulation and inflammation[27]. Genes such as *DPYSL5* and *MAPT* play crucial roles in nervous system development and are associated with neurodegenerative diseases like AD, potentially affecting CSF dynamics and glymphatic clearance[28, 29]. *GCAT* participates in amino acid metabolism[30], while *KHK* plays a key role in fructose metabolism[31], and *GNA12* regulates cellular metabolism via the *TOR* signaling pathway[32]. such as *TRIOBP* and *SNAP29*, likely influence CSF flow and waste clearance. Additionally, some genes, including *C16orf95* and *ABHD1*, may regulate molecular exchange between CSF and the lymphatic system, warranting further investigation.

Gene enrichment analysis using the DAVID platform revealed that the candidate genes are significantly enriched in pathways related to neuronal development, cell structure organization, signal transduction, enzyme activity, and metabolic processes (Fig. 4b and Supplementary Table 11). The single-cell disease relevance score (scDRS) algorithm showed significant enrichment of these genes in astrocytes (r = 0.11, p < 0.001) and oligodendrocytes (r = 0.12, p < 0.001) (Fig. 4c-d). Furthermore, the GWAS Catalog associated these genes with various brain imaging metrics, psychological and behavioral traits, neurological diseases such as Parkinson’s and Alzheimer’s, metabolic and blood indicators, lifestyle factors, and molecular markers, highlighting their potential roles in neural health and related biological processes (Supplementary Table 12).

Exploratory analyses identified enrichment of ALPS index-related genes in validated drug targets for other indications. For example, *CRHR1* targets include Flortaucipir F-18 and PBT-1033 used in diagnosing Cushing’s syndrome and psychiatric disorders, respectively[33, 34]. *MAPT* targets like Paclitaxel and Docetaxel are utilized in cancer treatment and Alzheimer’s research[35, 36]. *LPAR1*-driven drug BMS-986020 is currently in clinical trials [37], while *SERPIND1* and *TNIK*-driven drugs such as Sulodexide, Bemiparin, and Fostamatinib are used for treating venous thrombosis, rheumatoid arthritis, and immune thrombocytopenia[38, 39] (Supplementary Table 13). These findings suggest that the identified candidate genes not only play crucial roles in the biological mechanisms underlying the ALPS index but also represent potential targets for therapeutic interventions.

### Association Analysis Between ALPS Index and Five Major Categories of Non-Imaging Phenotypes

Previous small-scale studies have linked the ALPS index to diseases such as dementia, cerebrovascular disease, Parkinson’s, sleep disorders, type 2 diabetes, migraines, epilepsy, schizophrenia, multiple sclerosis, neuromyelitis optica, and glioma. To systematically investigate these associations, we analyzed the rank-inverse normal transformed ALPS index from 36,997 participants against 2,121 non-imaging phenotypes from the UK Biobank, categorized into Biomarkers and Physical Measurements, Medical Conditions and Treatments, Cognitive and Mental Health, Lifestyle and Social Factors, and Sex-specific Factors (Supplementary Table 14). After adjusting for age, sex, and assessment center, 181 phenotypes showed significant associations (β range: −0.675 to 0.435; p_FDR_: 1.44□×□10□²□ to 4.99□×□10□²) (Fig. 5 and Supplementary Table 15). Complete results for all 2,121 phenotypes are detailed in Supplementary Table 16.

**Figure 5.**
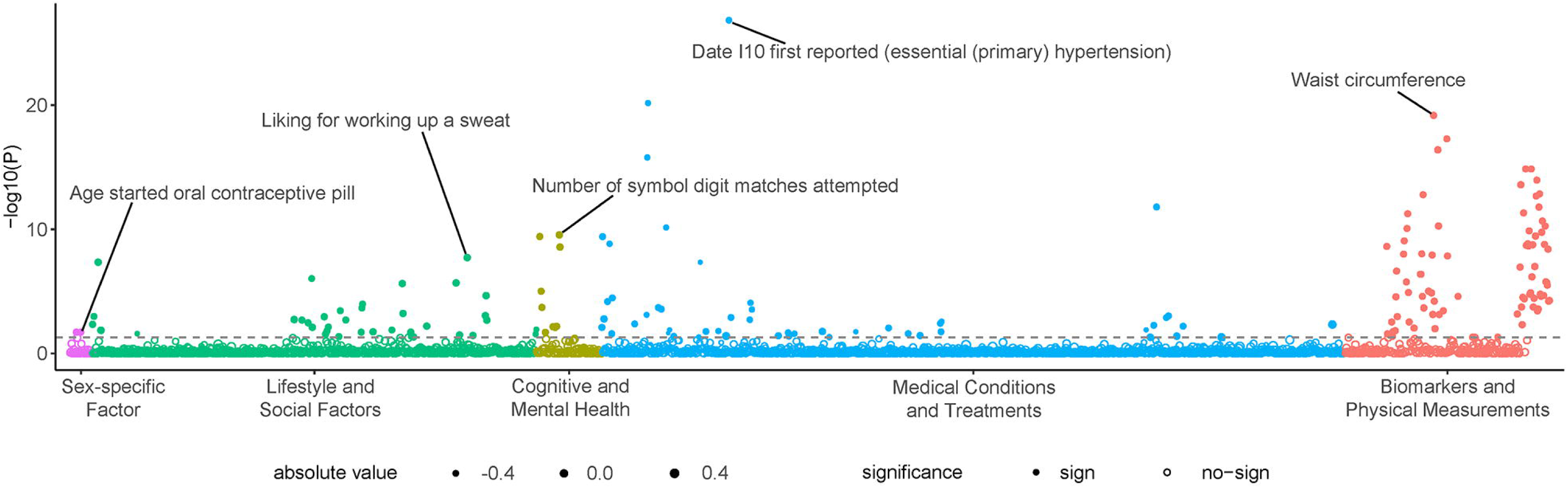
Manhattan plot showing associations of phenotypes with ALPS Index, grouped by categories. The x-axis represents phenotypes, and the y-axis represents the −log10 of uncorrected p values of two-sided test for linear regression between each phenotype and the ALPS Index (see Supplementary Table 14 for detailed results). Each dot represents one phenotype, and the colours indicate their according categories. The size of the dots corresponds to the magnitude (absolute value) of the effect between the phenotype and ALPS Index. The solid dots represent phenotypes that exhibited significant associations with the ALPS Index. The dashed lines indicate the threshold to survive Benjamini-Hochberg procedure (FDR correction).

Significant associations were observed in key categories, particularly blood cells, blood pressure, biochemistry, and body measurements. Blood cell-related phenotypes, including red and white blood cell counts, reticulocyte count, etc., displayed negative associations with the ALPS index (β = −0.042 to −0.014, p_FDR_ = 1.09 × 10□³ to 2.64 × 10□²), suggesting that alterations in blood composition may affect cerebrovascular health and brain fluid circulation. Blood pressure indicators (diastolic and systolic) also exhibited negative associations (β = −0.051 to −0.033, p_FDR_ = 5.14 × 10□¹□ to 1.41 × 10□□), indicating that hypertension may impede cerebrovascular function and glymphatic clearance. Body measurements, including weight, BMI, and waist circumference, were significantly negatively correlated with the ALPS index (β = −0.047 to −0.015, p_FDR_ = 1.09 × 10□¹□ to 6.67 × 10□²□), while height showed a positive association (β = 0.013 to 0.016, p_FDR_ = 1.01 × 10□² to 6.64 × 10□□), suggesting that changes in body fat distribution may decrease glymphatic function, whereas healthier body composition may enhance brain fluid circulation. Additionally, fat mass indicators (whole-body, trunk, and leg fat mass) exhibited negative associations with the ALPS index (β = −0.039 to −0.015, p_FDR_ = 1.09 × 10□¹□ to 1.34 × 10□¹□), implying that excessive fat may inhibit fluid flow. Lean mass in the arms and legs was also negatively correlated (β = −0.019 to −0.017, p_FDR_ = 1.67 × 10□□ to 3.75 × 10□□), suggesting that muscle loss may disrupt fluid balance. Biochemical markers such as uric acid, glycated haemoglobin, and C-reactive protein displayed significant negative associations with the ALPS index (β = −0.056 to −0.029, p_FDR_ = 1.24 × 10□□ to 4.18 × 10□□), indicating that metabolic abnormalities may impair brain fluid balance and glymphatic function. Finally, additional indicators from pulmonary function, ocular measurements, and impedance measures were assessed. Pulmonary function metrics (Forced Vital Capacity [FVC] and Forced Expiratory Volume in 1 second [FEV□]) positively correlated with the ALPS index, suggesting respiratory health supports brain fluid balance. Additionally, eye surgeries and impedance measures may reflect changes in brain fluid dynamics.

Diabetes, including non-insulin-dependent and insulin-dependent types, obesity, and dyslipidemia were significantly negatively correlated with the ALPS index (β = −0.286 to −0.058, p_FDR_ = 1.48 × 10□□ to 2.65 × 10□□), suggesting that metabolic abnormalities may disrupt brain fluid dynamics via vascular and metabolic pathways. Cardiovascular and cerebrovascular diseases, such as hypertension, angina, and cerebral infarction, also showed negative associations (β = −0.240 to −0.101, p_FDR_ = 1.26 × 10□³ to 2.89 × 10□□), indicating impaired cerebrovascular function and glymphatic clearance. Medical interventions, including antihypertensive medications and aspirin, also correlated negatively with the ALPS index (β = −0.122 to −0.088, p_FDR_ = 9.46 × 10□□ to 1.30 × 10□³), potentially affecting glymphatic clearance through metabolic and vascular regulation. Neurological disorders, including MS, AD, and dementia, exhibited significant negative associations (β = −0.592 to −0.506, p_FDR_ = 1.27 × 10□² to 2.36 × 10□²), suggesting impaired neuronal function and waste clearance. Kidney diseases (acute and chronic renal failure) and respiratory diseases (asthma and Chronic Obstructive Pulmonary Disease) were negatively correlated with the ALPS index, suggesting that impaired renal and lung function may disrupt brain fluid balance, whereas bilateral vasectomy showed a positive association, potentially related to metabolic or vascular changes.

In the cognitive and mental health domain, the ALPS index was positively correlated with the number of attempts and correct matches in symbol-matching tasks and negatively correlated with task completion and identification times (β = −0.039 to 0.050, p_FDR_ = 2.02□×□10□² to 2.85□×□10□¹□), indicating that efficient brain fluid circulation is associated with enhanced cognitive performance. In health behaviours and lifestyle factors, physical exercise indicators positively correlated with the ALPS index (β = 0.033 to 0.067, p_FDR_ = 1.91□×□10□□ to 4.01□×□10□²), suggesting enhanced brain fluid circulation, whereas smoking, increased computer use, video gaming, and circadian rhythm disturbances negatively correlated (β = −0.131 to −0.036, p_FDR_ = 1.95□×□10□² to 3.41□×□10□³), indicating impaired glymphatic clearance. Additionally, socioeconomic factors and birth weight showed positive associations (β = 0.040 to 0.051, p_FDR_ = 2.05□×□10□³ to 4.50□×□10□□), reflecting better health management, while sex-specific factors such as number of children and age at first use of oral contraceptives were also associated with the ALPS index, suggesting potential sex-specific influences on brain fluid dynamics.

### Polygenic Risk Score Validation of ALPS Index Associations Across Five Phenotype Categories

In the validation process, to evaluate the predictive efficacy of the polygenic risk score (PRS)[24] for the ALPS Index, we calculated the PRS in 370,920 UKBB participants without directly measured ALPS Index data, using PRS-CS and GWAS summary statistics from 36,111 White British participants. The ALPS index-PRS was set as the independent variable, with age, sex, genotyping array, top 10 genetic principal components, and assessment centre as covariates. A positive association was observed, indicating strong predictive capacity (Supplementary Fig. 4). PRS-PheWAS analysis assessed 181 phenotypes, revealing significant associations with 42 phenotypes, with consistent effect directions for 29 phenotypes (Supplementary Tables 15 and 16). Significant associations were found for 7.5% of biomarkers and physical measurements, 0.8% of lifestyle factors, and 0.2% of medical conditions. Negative associations with the ALPS Index were observed for diastolic and systolic blood pressure, haematological markers (red blood cell distribution width and reticulocyte count), the date of first AD diagnosis, time spent playing computer games, and dietary diversity. In contrast, positive associations were identified for lung function indicators (FVC and FEV□), body measurements (height, Sex Hormone-Binding Globulin, vitamin D levels), birth weight, walking speed, preference for green olives, and the age at first use of glasses or contact lenses.

### Validation of ALPS Index Associations with Neurological Diseases Using Mendelian Randomization and Cox Regression

In the association analysis, significant negative associations were identified between the ALPS Index and neurological diseases, including AD, unspecified dementia, MS, cerebral infarction, stroke (unspecified as haemorrhage or infarction), and other cerebrovascular diseases (β = −0.592 to −0.206, p_FDR_ = 4.50□×□10□^8^ to 4.77□×□10^−2^). To further validate these associations, Cox proportional hazards models were applied to evaluate time-to-event data, providing insights into the influence of the ALPS Index on disease risk or progression over time. The ALPS Index was divided into three categories based on tertiles: low, medium, and high. Compared to the low ALPS Index category, stronger protective effects were observed, particularly for AD (HR = 0.645, p = 6.09 × 10□□) and MS (HR = 0.655, p = 1.34 × 10□□), indicating a significant reduction in disease risk (Fig. 6 and Supplementary Table 17). Following this, Mendelian randomization (MR)[25] was employed to assess potential causal relationships between the ALPS Index and AD and ischemic stroke, showing a protective effect (OR = 0.889 for AD, p = 2.630□×□10^−2^; OR = 0.867 for ischemic stroke, p = 2.590□×□10^−2^, Supplementary Table 18). MR-Egger intercept tests indicated no horizontal pleiotropy and heterogeneity tests confirmed these associations (Supplementary Table 19). Collectively, these findings indicate that a higher ALPS Index is associated with a reduced risk of AD and MS, as shown by association and Cox regression analyses, as well as a reduced risk of AD and ischemic stroke, as supported by MR analysis.

**Figure 6.**
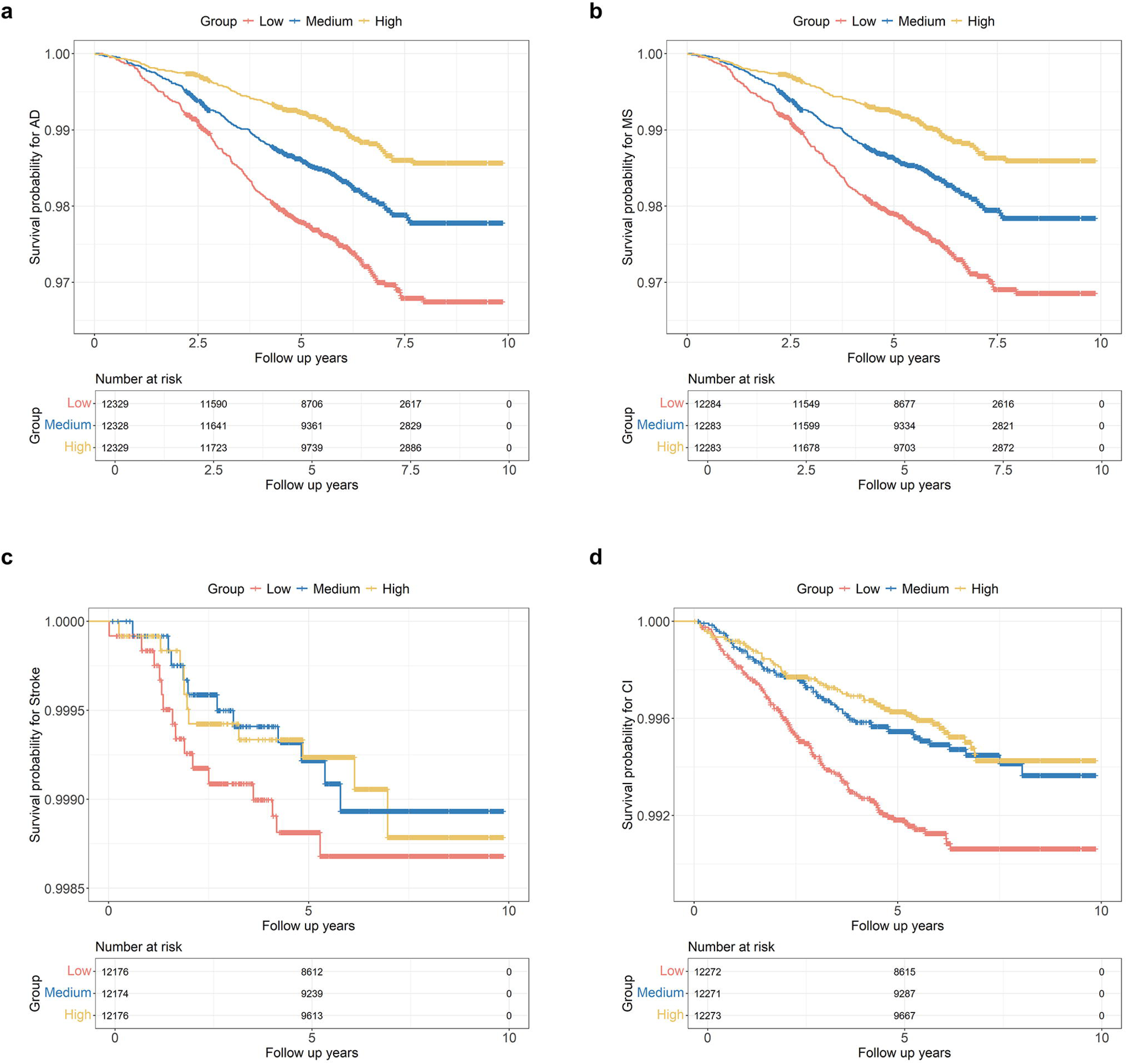
Cox Proportional Hazards Analysis of ALPS Index and Neurological Diseases. This figure shows the Cox proportional hazards model results for the association between the ALPS Index and four common neurological diseases: AD, MS, cerebral infarction (ischemic stroke, IS), and stroke (unspecified as hemorrhage or infarction). The curves represent the time-to-event analysis, with the x-axis indicating the follow-up time in years and the y-axis showing the survival probability for the respective neurological diseases. Three categories of the ALPS Index were analyzed: continuous, medium, and high. The hazard ratios (HR) for each disease and category are provided along with their corresponding 95% confidence intervals (L95, U95) and p-values, as adjusted for age, sex, genotyping array, Townsend deprivation index, and body mass index (BMI). Participants with pre-existing neurological conditions were excluded from the analysis.

## Discussion

To our knowledge, this study represents the largest individual-level GWAS to date investigating the genetic architecture of the ALPS index, a key indicator of glymphatic system function within the brain. Utilizing data from 36,997 participants in the UKBB, we employed a novel analytical framework to calculate the ALPS index from DTI data and conducted comprehensive genetic analyses. Our GWAS identified 14 independent genome-wide significant loci associated with the ALPS index, encompassing 3,814 significant single nucleotide polymorphisms (SNPs). The candidate genes implicated by these loci are involved in critical biological pathways related to neuronal development, vascular integrity, and metabolic processes. Furthermore, our extensive association analysis revealed significant associations between the ALPS index and a diverse range of non-imaging phenotypes across five major categories: biomarkers and physical measurements, medical conditions and treatments, cognitive and mental health, lifestyle and social factors, and sex-specific factors. These findings underscore the genetic basis of glymphatic function and highlight its potential role in the risk stratification and early detection of neurodegenerative diseases. Collectively, our results provide valuable insights into the genetic determinants of brain fluid dynamics and their implications for neural health.

A total of 3,814 variants were identified across 14 genome-wide significant loci. Analysis of these loci associated with the ALPS index highlighted their enrichment in intronic and intergenic regions, indicating a potential regulatory role in glymphatic function. Notably, only 1.34% of the variants were exonic, suggesting that non-coding regions may significantly influence glymphatic activity and brain health. This conclusion is further supported by the presence of variants in regions of active chromatin, including open chromatin areas, suggesting involvement in the transcriptional regulation of genes related to glymphatic efficiency. These risk variants also showed enrichment in genetic regions linked to traits such as white matter integrity, fibre tract connectivity, brain structure volume, inflammation, and metabolism, highlighting the interplay between glymphatic function and broader neurobiological processes. Furthermore, results from LDSC regression indicated that the observed genomic inflation was primarily due to polygenicity rather than population stratification, supporting the robustness of the findings. Additional analyses using GCTA-COJO identified 15 independent SNPs within the 14 significant loci identified by FUMA, further validating these associations. Collectively, these findings suggest that genetic variants influencing the ALPS index predominantly exert regulatory effects, with broader implications for understanding the genetic basis of glymphatic function and its link to neurological health and disease risk.

The identified variations associated with the ALPS Index were primarily located in gene regions implicated in regulating white matter integrity, fibre tract connectivity, and metabolic function. These aspects are closely linked to the mechanisms underlying brain waste clearance[6]. Functional annotation of the ALPS Index-associated loci revealed key genes related to cerebrovascular function, including *GNA12*, *SERPIND1*, and *LPAR1*, which may enhance the efficiency of the glymphatic system by regulating brain fluid flow[40]. Notably, *GNA12* modulates smooth muscle contraction through the PI3K/ROCK signalling pathway and protects endothelial cells by maintaining miR-155 levels, underscoring its pivotal role in vascular function regulation[41]. *GNA12* was consistently identified in both GWAS and functional annotation analyses as a gene linked to the regulation of brain fluid dynamics, particularly in regions associated with smooth muscle and cerebrovascular function. This suggests that its variants may impact glymphatic waste clearance by altering cerebral hemodynamics[42].

Additionally, our study revealed associations between the ALPS Index and genes implicated in neurodevelopment and neurodegenerative diseases. Notably, *MAPT* was frequently identified in analyses related to the ALPS Index[43]. *MAPT* encodes the Tau protein, whose abnormal accumulation is a hallmark of AD[44]. Our findings suggest that variants in *MAPT* may influence CSF clearance by promoting Tau protein accumulation, potentially accelerating the progression of neurodegenerative diseases. Furthermore, *MAPT*-targeted therapies, such as paclitaxel and docetaxel—commonly used in cancer treatment—underscore *MAPT*’s role in cytoskeletal dynamics and protein metabolism[45]. This indicates that Tau accumulation, in addition to its connection with neuronal damage, may also impair glymphatic clearance, thereby increasing the risk of neurodegenerative diseases.

Among the candidate genes, *LPAR1* emerged as particularly significant. *LPAR1* is integral to cell migration and morphogenesis and plays a critical role in myelination and functional connectivity within the brain[46]. Our findings indicate that *LPAR1* is closely linked to the regulation of glymphatic function, particularly in relation to brain waste clearance.[47]. Moreover, drugs targeting *LPAR1*, such as BMS-986020, which are currently undergoing clinical trials, present promising avenues for future research into therapies aimed at addressing glymphatic system dysfunction[37].

Another significant gene, *DPYSL5*, was repeatedly associated with glymphatic system function, particularly in neuronal migration and axon guidance[28]. *DPYSL5* plays a crucial role in regulating brain fluid dynamics and waste clearance, with mutations potentially impacting the ALPS Index and, consequently, neurodevelopment and cognitive function. In our exome-wide analysis, the synonymous variant (p.Gly558Gly) of *DPYSL5* was significantly associated with the ALPS Index, further supporting its involvement in glymphatic regulation. Given its critical function during neurodevelopment, variations in *DPYSL5* may indirectly influence glymphatic waste clearance by modulating neuronal migration and synaptic transmission[48].

*SERPIND1* was identified in multiple analyses related to the ALPS Index as a gene closely tied to CSF clearance. It primarily regulates blood coagulation and inflammatory responses, thereby influencing vascular function and brain fluid dynamics[49]. Drugs targeting *SERPIND1*, like sulodexide and heparin, which are already used for venous thrombosis prevention and treatment, support its potential as a therapeutic target for regulating glymphatic function[38]. Additionally, *TNIK* modulates neurodevelopment and synaptic transmission, indirectly impacting CSF flow and metabolic waste clearance[50]. Current drugs targeting *TNIK*, such as fostamatinib, used for rheumatoid arthritis and immune thrombocytopenia, suggest that this gene could serve as a novel target for future therapies addressing glymphatic system dysfunction[51].

The gene enrichment analyses revealed that candidate genes associated with the ALPS index are involved in critical neuronal and cellular processes, such as neuronal development and metabolic pathways. This aligns with existing research on the genetic underpinnings of neurodevelopmental and neurodegenerative disorders. For example, genes related to neuronal structure and signal transduction pathways have been previously implicated in neurodegenerative conditions like AD and PD[52]. The scDRS algorithm’s identification of significant gene enrichment in astrocytes and oligodendrocytes underscores their role in neural homeostasis and disease progression, consistent with studies highlighting astrocytes’ involvement in neuroinflammation and myelination processes, which are crucial for maintaining brain health[53]. Furthermore, the significant associations between candidate genes and brain imaging metrics, cognitive traits, and metabolic indicators point to a complex genetic architecture underlying cognitive and psychological traits, reinforcing the genetic overlap between neurological and metabolic conditions[54]. Additionally, drug target enrichment analyses suggest a potential for repurposing existing drugs that target ALPS-associated genes, such as *CRHR1*- and *MAPT*-driven therapies, for neurodegenerative conditions. These findings highlight the translational potential of ALPS-related genes in guiding therapeutic strategies for both neurological and non-neurological conditions.

Our study found significant negative associations between the ALPS index and various haematological traits, including red blood cell count, white blood cell count, and reticulocyte count. This suggests that alterations in immune function and blood composition may profoundly affect brain health by disrupting fluid balance and waste clearance[55]. Furthermore, the negative associations with diastolic and systolic blood pressure reinforce the detrimental effects of hypertension on cerebrovascular health, indicating that elevated blood pressure may hinder CSF flow and reduce waste clearance efficiency. Previous studies have linked hypertension to white matter lesions and disturbances in CSF circulation, supporting these findings[56].

Anthropometric measures such as weight, BMI, and waist circumference showed negative associations with the ALPS Index, while height correlated positively, suggesting that increased adiposity may impair glymphatic function by disrupting brain fluid dynamics[57, 58]. Negative associations with biochemical markers like uric acid, glycated haemoglobin, and C-reactive protein further link metabolic abnormalities and chronic inflammation to impaired brain waste clearance[59]. Medical conditions related to metabolic dysfunction—including diabetes and its subtypes, obesity, and dysregulated lipid metabolism—were also negatively associated with the ALPS Index. These findings indicate that metabolic dysregulation, especially chronic fluctuations in blood glucose and increased adiposity, may impair glymphatic clearance by affecting the cerebrovascular system and altering CSF composition and flow[60].

In the cardiovascular domain, significant negative associations were observed between hypertension, angina, and the ALPS Index, reinforcing the known link between hypertension and cerebrovascular dysfunction. Elevated blood pressure may compromise vascular structure and elasticity, impeding normal CSF flow and reducing waste clearance efficiency[56]. Chronic hypertension has been shown to contribute to white matter lesions and abnormal CSF circulation, adversely affecting brain health[61]. Additionally, ischemic stroke and cerebrovascular diseases, such as cerebral infarction, were significantly correlated with lower ALPS Index values, indicating that disruptions in cerebral blood supply during stroke can severely impair CSF flow and glymphatic clearance[62], potentially exacerbating neuronal damage and neurodegeneration during recovery[63].

Neurological diseases, including AD, MS, and unspecified dementia, showed significant negative associations with the ALPS Index, implying that these conditions may hinder glymphatic clearance by disrupting immune and metabolic functions in the brain. Patients with these conditions frequently exhibit restricted CSF flow and accumulated metabolic waste, both of which are closely linked to impaired glymphatic function[64]. Moreover, the inflammatory responses and immune dysregulation associated with neurodegenerative diseases, particularly in MS, may further exacerbate these impairments, highlighting the intricate relationship between glymphatic efficiency and neurological health[65]. In addition, validation through Cox regression confirmed a protective role of a higher ALPS Index in reducing the risk of AD and MS, while MR supported its protective effect on AD and ischemic stroke (cerebral infarction). These findings further reinforce the hypothesis that improved glymphatic function, as reflected by a higher ALPS Index, is associated with a lower risk of neurodegenerative and cerebrovascular conditions. This underscores the potential of the ALPS Index as a valuable marker for assessing both glymphatic function and neurological disease risk.

Although this study demonstrates a strong capacity for discovery, several limitations should be noted. This study was primarily conducted in populations of European ancestry, validation in other ethnic groups is a crucial step to ensure the generalizability of the findings. Future research should also explore therapeutic interventions targeting the glymphatic system, especially in individuals carrying rare loss-of-function variants. Moreover, integrating multi-omics data (e.g., proteomics, metabolomics) may help uncover the molecular mechanisms underlying glymphatic system function.

## Conclusion

In conclusion, this study systematically elucidates the genetic architecture of the ALPS Index, identifying key loci and pathways associated with glymphatic system function and its broad impact on brain health. These findings provide new insights into the role of the glymphatic system in neurodegenerative diseases and offer a foundation for the development of therapeutic strategies targeting glymphatic dysfunction.

## Abbreviations

AD: Alzheimer’s disease
CADD: Combined Annotation Dependent Depletion
CSF: Cerebrospinal fluid
DTI: Diffusion tensor imaging
DTI-ALPS: Diffusion Tensor Imaging Along the Perivascular Space
DWI: Diffusion-weighted imaging
eQTL: Expression Quantitative Trait Loci
FDR: False discovery rate
FEV□: Forced Expiratory Volume in 1 second
FVC: Forced Vital Capacity
GCTA-COJO: Conditional and joint multiple-SNP analysis using Genome-wide Complex Trait Analysis
GO: Gene Ontology
GTEx v8: Genotype-Tissue Expression project version 8
GWAS: Genome-wide association study
IVW: Inverse-variance weighted
LDSC: Linkage Disequilibrium SCore regression
MAF: Minor Allele Frequency
MAGMA: Multi-marker Analysis of GenoMic Annotation
MR: Mendelian Randomization
MS: Multiple sclerosis
OR: Odds ratio
PRS: Polygenic risk score
ROIs: Regions of interest
scDRS: Single-cell disease relevance score
SNP: Single nucleotide polymorphism
SNVs: Single nucleotide variants
sQTL: Splicing Quantitative Trait Loci
TWAS: Transcriptome-wide association study
UKBB: UK Biobank

## Declarations

### Consent for publication

All authors have read and agreed to the published version of the manuscript.

### Availability of data and materials

Data from the UK Biobank Diffusion-Weighted Imaging (DWI) scans were collected according to the published protocol (https://biobank.ctsu.ox.ac.uk/crystal/refer.cgi?id=2367). Permission to use the UK Biobank Resource was obtained via material transfer agreement as part of Data Access Application 221671. All imaging data, phenotypes and genetics data are made available by UK Biobank via their standard data access procedure (see http://www.ukbiobank.ac.uk/register-apply). All other data supporting the findings of this study are available within the article, the supplementary information or the supplementary data files.

### Authors’ contributions

C.S. conceived and designed this study. D.M. drafted the manuscript. C.S., S.T., C.M., Y.J., Y.X., Y.W., S.S., and O.A.A. revised the manuscript. D.M., S.L., C.Z., Z.W., Y.S., S.Q., Y.L., C.H., Y.F., X.H., M.L., and R.Y. performed statistical analysis. C.S. supervised the project. C.S. and C.M. obtained funding. All authors have read and approved the final version of the manuscript.

## Acknowledgements

This research was conducted using the UKBB Resource under application number 221671. We are grateful to UK Biobank for making the data available and to all UK Biobank study participants who generously donated their time to make this resource possible. The funders had no role in study design, data collection and analysis, decision to publish or preparation of the manuscript.

## References

1. Lohela TJ, Lilius TO, Nedergaard M: The glymphatic system: implications for drugs for central nervous system diseases. Nature reviews Drug discovery 2022, 21(10):763–779.

2. Klarica M, Radoš M, Orešković D: The Movement of Cerebrospinal Fluid and Its Relationship with Substances Behavior in Cerebrospinal and Interstitial Fluid. Neuroscience 2019, 414:28–48.

3. Kazemi H, Johnson DC: Regulation of cerebrospinal fluid acid-base balance. Physiological reviews 1986, 66(4):953–1037.

4. Wood KH, Nenert R, Miften AM, Kent GW, Sleyster M, Memon RA, Joop A, Pilkington J, Memon AA, Wilson RN et al: Diffusion Tensor Imaging-Along the Perivascular-Space Index Is Associated with Disease Progression in Parkinson’s Disease. Mov Disord 2024.

5. Taoka T, Ito R, Nakamichi R, Kamagata K, Sakai M, Kawai H, Nakane T, Abe T, Ichikawa K, Kikuta J et al: Reproducibility of diffusion tensor image analysis along the perivascular space (DTI-ALPS) for evaluating interstitial fluid diffusivity and glymphatic function: CHanges in Alps index on Multiple conditiON acquIsition eXperiment (CHAMONIX) study. Japanese journal of radiology 2022, 40(2):147–158.

6. Gouveia-Freitas K, Bastos-Leite AJ: Perivascular spaces and brain waste clearance systems: relevance for neurodegenerative and cerebrovascular pathology. Neuroradiology 2021, 63(10):1581–1597.

7. Alfaro-Almagro F, Jenkinson M, Bangerter NK, Andersson JLR, Griffanti L, Douaud G, Sotiropoulos SN, Jbabdi S, Hernandez-Fernandez M, Vallee E et al: Image processing and Quality Control for the first 10,000 brain imaging datasets from UK Biobank. NeuroImage 2018, 166:400–424.

8. Liu X, Barisano G, Shao X, Jann K, Ringman JM, Lu H, Arfanakis K, Caprihan A, DeCarli C, Gold BT et al: Cross-Vendor Test-Retest Validation of Diffusion Tensor Image Analysis along the Perivascular Space (DTI-ALPS) for Evaluating Glymphatic System Function. Aging and disease 2024, 15(4):1885–1898.

9. Mbatchou J, Barnard L, Backman J, Marcketta A, Kosmicki JA, Ziyatdinov A, Benner C, O’Dushlaine C, Barber M, Boutkov B et al: Computationally efficient whole-genome regression for quantitative and binary traits. Nature genetics 2021, 53(7):1097–1103.

10. Bulik-Sullivan BK, Loh PR, Finucane HK, Ripke S, Yang J, Patterson N, Daly MJ, Price AL, Neale BM: LD Score regression distinguishes confounding from polygenicity in genome-wide association studies. Nature genetics 2015, 47(3):291–295.

11. Watanabe K, Taskesen E, van Bochoven A, Posthuma D: Functional mapping and annotation of genetic associations with FUMA. Nature communications 2017, 8(1):1826.

12. Yang J, Ferreira T, Morris AP, Medland SE, Madden PA, Heath AC, Martin NG, Montgomery GW, Weedon MN, Loos RJ et al: Conditional and joint multiple-SNP analysis of GWAS summary statistics identifies additional variants influencing complex traits. Nature genetics 2012, 44(4):369–375, s361-363.

13. de Leeuw CA, Mooij JM, Heskes T, Posthuma D: MAGMA: generalized gene-set analysis of GWAS data. PLoS computational biology 2015, 11(4):e1004219.

14. Wang K, Li M, Hakonarson H: ANNOVAR: functional annotation of genetic variants from high-throughput sequencing data. Nucleic acids research 2010, 38(16):e164.

15. Gusev A, Ko A, Shi H, Bhatia G, Chung W, Penninx BW, Jansen R, de Geus EJ, Boomsma DI, Wright FA et al: Integrative approaches for large-scale transcriptome-wide association studies. Nature genetics 2016, 48(3):245–252.

16. Giambartolomei C, Vukcevic D, Schadt EE, Franke L, Hingorani AD, Wallace C, Plagnol V: Bayesian test for colocalisation between pairs of genetic association studies using summary statistics. PLoS genetics 2014, 10(5):e1004383.

17. Sherman BT, Hao M, Qiu J, Jiao X, Baseler MW, Lane HC, Imamichi T, Chang W: DAVID: a web server for functional enrichment analysis and functional annotation of gene lists (2021 update). Nucleic acids research 2022, 50(W1):W216–w221.

18. Zhang MJ, Hou K, Dey KK, Sakaue S, Jagadeesh KA, Weinand K, Taychameekiatchai A, Rao P, Pisco AO, Zou J et al: Polygenic enrichment distinguishes disease associations of individual cells in single-cell RNA-seq data. Nature genetics 2022, 54(10):1572–1580.

19. Xie Z, Bailey A, Kuleshov MV, Clarke DJB, Evangelista JE, Jenkins SL, Lachmann A, Wojciechowicz ML, Kropiwnicki E, Jagodnik KM et al: Gene Set Knowledge Discovery with Enrichr. Current protocols 2021, 1(3):e90.

20. Cannon M, Stevenson J, Stahl K, Basu R, Coffman A, Kiwala S, McMichael JF, Kuzma K, Morrissey D, Cotto K et al: DGIdb 5.0: rebuilding the drug-gene interaction database for precision medicine and drug discovery platforms. Nucleic acids research 2024, 52(D1):D1227–d1235.

21. Finan C, Gaulton A, Kruger FA, Lumbers RT, Shah T, Engmann J, Galver L, Kelley R, Karlsson A, Santos R et al: The druggable genome and support for target identification and validation in drug development. Science translational medicine 2017, 9(383).

22. Knox C, Wilson M, Klinger CM, Franklin M, Oler E, Wilson A, Pon A, Cox J, Chin NEL, Strawbridge SA et al: DrugBank 6.0: the DrugBank Knowledgebase for 2024. Nucleic acids research 2024, 52(D1):D1265–d1275.

23. Millard LAC, Davies NM, Gaunt TR, Davey Smith G, Tilling K: Software Application Profile: PHESANT: a tool for performing automated phenome scans in UK Biobank. International journal of epidemiology 2018, 47(1):29–35.

24. Ge T, Chen CY, Ni Y, Feng YA, Smoller JW: Polygenic prediction via Bayesian regression and continuous shrinkage priors. Nature communications 2019, 10(1):1776.

25. Hemani G, Zheng J, Elsworth B, Wade KH, Haberland V, Baird D, Laurin C, Burgess S, Bowden J, Langdon R et al: The MR-Base platform supports systematic causal inference across the human phenome. eLife 2018, 7.

26. Andreeva AV, Kutuzov MA, Vaiskunaite R, Profirovic J, Meigs TE, Predescu S, Malik AB, Voyno-Yasenetskaya T: G alpha12 interaction with alphaSNAP induces VE-cadherin localization at endothelial junctions and regulates barrier function. The Journal of biological chemistry 2005, 280(34):30376–30383.

27. Nourse J, Braun J, Lackner K, Hüttelmaier S, Danckwardt S: Large-scale identification of functional microRNA targeting reveals cooperative regulation of the hemostatic system. Journal of thrombosis and haemostasis : JTH 2018, 16(11):2233–2245.

28. Jeanne M, Demory H, Moutal A, Vuillaume ML, Blesson S, Thépault RA, Marouillat S, Halewa J, Maas SM, Motazacker MM et al: Missense variants in DPYSL5 cause a neurodevelopmental disorder with corpus callosum agenesis and cerebellar abnormalities. Am J Hum Genet 2021, 108(5):951–961.

29. Yoshida H, Goedert M: Phosphorylation of microtubule-associated protein tau by AMPK-related kinases. Journal of neurochemistry 2012, 120(1):165–176.

30. Liu YJ, Janssens GE, McIntyre RL, Molenaars M, Kamble R, Gao AW, Jongejan A, Weeghel MV, MacInnes AW, Houtkooper RH: Glycine promotes longevity in Caenorhabditis elegans in a methionine cycle-dependent fashion. PLoS Genet 2019, 15(3):e1007633.

31. Patel C, Douard V, Yu S, Tharabenjasin P, Gao N, Ferraris RP: Fructose-induced increases in expression of intestinal fructolytic and gluconeogenic genes are regulated by GLUT5 and KHK. American journal of physiology Regulatory, integrative and comparative physiology 2015, 309(5):R499–509.

32. Gismondi A, Nanni V, Monteleone V, Colao C, Di Marco G, Canini A: Plant miR171 modulates mTOR pathway in HEK293 cells by targeting GNA12. Molecular biology reports 2021, 48(1):435–449.

33. Torre BG, Albericio F: The Pharmaceutical Industry in 2020. An Analysis of FDA Drug Approvals from the Perspective of Molecules. *Molecules (Basel*, Switzerland) 2021, 26(3).

34. Coric V, Feldman HH, Oren DA, Shekhar A, Pultz J, Dockens RC, Wu X, Gentile KA, Huang SP, Emison E et al: Multicenter, randomized, double-blind, active comparator and placebo-controlled trial of a corticotropin-releasing factor receptor-1 antagonist in generalized anxiety disorder. Depression and anxiety 2010, 27(5):417–425.

35. Saloustros E, Mavroudis D, Georgoulias V: Paclitaxel and docetaxel in the treatment of breast cancer. Expert opinion on pharmacotherapy 2008, 9(15):2603–2616.

36. Pontecorvo MJ, Devous MD, Kennedy I, Navitsky M, Lu M, Galante N, Salloway S, Doraiswamy PM, Southekal S, Arora AK et al: A multicentre longitudinal study of flortaucipir (18F) in normal ageing, mild cognitive impairment and Alzheimer’s disease dementia. Brain 2019, 142(6):1723–1735.

37. Palmer SM, Snyder L, Todd JL, Soule B, Christian R, Anstrom K, Luo Y, Gagnon R, Rosen G: Randomized, Double-Blind, Placebo-Controlled, Phase 2 Trial of BMS-986020, a Lysophosphatidic Acid Receptor Antagonist for the Treatment of Idiopathic Pulmonary Fibrosis. Chest 2018, 154(5):1061–1069.

38. Wang P, Chi L, Zhang Z, Zhao H, Zhang F, Linhardt RJ: Heparin: An old drug for new clinical applications. Carbohydrate polymers 2022, 295:119818.

39. Kang Y, Jiang X, Qin D, Wang L, Yang J, Wu A, Huang F, Ye Y, Wu J: Efficacy and Safety of Multiple Dosages of Fostamatinib in Adult Patients With Rheumatoid Arthritis: A Systematic Review and Meta-Analysis. Frontiers in pharmacology 2019, 10:897.

40. Räsänen J, Heikkinen S, Mäklin K, Lipponen A, Kuulasmaa T, Mehtonen J, Korhonen VE, Junkkari A, Grenier-Boley B, Bellenguez C et al: Risk Variants Associated With Normal Pressure Hydrocephalus: Genome-Wide Association Study in the FinnGen Cohort. Neurology 2024, 103(5):e209694.

41. Yoo EJ, Cao G, Koziol-White CJ, Ojiaku CA, Sunder K, Jude JA, Michael JV, Lam H, Pushkarsky I, Damoiseaux R et al: Gα(12) facilitates shortening in human airway smooth muscle by modulating phosphoinositide 3-kinase-mediated activation in a RhoA-dependent manner. British journal of pharmacology 2017, 174(23):4383–4395.

42. Vojinovic D, Adams HH, Jian X, Yang Q, Smith AV, Bis JC, Teumer A, Scholz M, Armstrong NJ, Hofer E et al: Genome-wide association study of 23,500 individuals identifies 7 loci associated with brain ventricular volume. Nat Commun 2018, 9(1):3945.

43. Hsu JL, Wei YC, Toh CH, Hsiao IT, Lin KJ, Yen TC, Liao MF, Ro LS: Magnetic Resonance Images Implicate That Glymphatic Alterations Mediate Cognitive Dysfunction in Alzheimer Disease. Annals of neurology 2023, 93(1):164–174.

44. Spina S, Schonhaut DR, Boeve BF, Seeley WW, Ossenkoppele R, O’Neil JP, Lazaris A, Rosen HJ, Boxer AL, Perry DC et al: Frontotemporal dementia with the V337M MAPT mutation: Tau-PET and pathology correlations. Neurology 2017, 88(8):758–766.

45. Ikeda H, Taira N, Hara F, Fujita T, Yamamoto H, Soh J, Toyooka S, Nogami T, Shien T, Doihara H et al: The estrogen receptor influences microtubule-associated protein tau (MAPT) expression and the selective estrogen receptor inhibitor fulvestrant downregulates MAPT and increases the sensitivity to taxane in breast cancer cells. Breast cancer research : BCR 2010, 12(3):R43.

46. Liu X, Pei M, Yu Y, Wang X, Gui J: Reduction of LPAR1 Expression in Neuroblastoma Promotes Tumor Cell Migration. Cancers 2022, 14(14).

47. Duperron MG, Knol MJ, Le Grand Q, Evans TE, Mishra A, Tsuchida A, Roshchupkin G, Konuma T, Trégouët DA, Romero JR et al: Genomics of perivascular space burden unravels early mechanisms of cerebral small vessel disease. Nature medicine 2023, 29(4):950–962.

48. Desprez F, Ung DC, Vourc’h P, Jeanne M, Laumonnier F: Contribution of the dihydropyrimidinase-like proteins family in synaptic physiology and in neurodevelopmental disorders. Frontiers in neuroscience 2023, 17:1154446.

49. Grover SP, Mackman N: Anticoagulant SERPINs: Endogenous Regulators of Hemostasis and Thrombosis. Frontiers in cardiovascular medicine 2022, 9:878199.

50. Coba MP, Komiyama NH, Nithianantharajah J, Kopanitsa MV, Indersmitten T, Skene NG, Tuck EJ, Fricker DG, Elsegood KA, Stanford LE et al: TNiK is required for postsynaptic and nuclear signaling pathways and cognitive function. J Neurosci 2012, 32(40):13987–13999.

51. Connell NT, Berliner N: Fostamatinib for the treatment of chronic immune thrombocytopenia. Blood 2019, 133(19):2027–2030.

52. Correction: Effectiveness of intraprocedural dual-phase cone-beam computed tomography in detecting hepatocellular carcinoma and improving treatment outcomes following conventional transarterial chemoembolization. PLoS One 2021, 16(2):e0247648.

53. Holtman IR, Bsibsi M, Gerritsen WH, Boddeke HW, Eggen BJ, van der Valk P, Kipp M, van Noort JM, Amor S: Identification of highly connected hub genes in the protective response program of human macrophages and microglia activated by alpha B-crystallin. Glia 2017, 65(3):460–473.

54. Spaepen E, Gunderson EA, Gibson D, Goldin-Meadow S, Levine SC: Meaning before order: Cardinal principle knowledge predicts improvement in understanding the successor principle and exact ordering. Cognition 2018, 180:59–81.

55. Liu J, Guo Y, Zhang C, Zeng Y, Luo Y, Wang G: Clearance Systems in the Brain, From Structure to Function. Frontiers in cellular neuroscience 2021, 15:729706.

56. Mestre H, Tithof J, Du T, Song W, Peng W, Sweeney AM, Olveda G, Thomas JH, Nedergaard M, Kelley DH: Flow of cerebrospinal fluid is driven by arterial pulsations and is reduced in hypertension. Nat Commun 2018, 9(1):4878.

57. Gray SM, Barrett EJ: Insulin transport into the brain. American journal of physiology Cell physiology 2018, 315(2):C125–c136.

58. Dadas A, Washington J, Janigro D: Cerebral Waste Accumulation and Glymphatic Clearance as Mechanisms of Human Neurological Diseases. Journal of neurology & neuromedicine 2016, 1(7):15–19.

59. Lu Y, Jiang X, Liu S, Li M: Changes in Cerebrospinal Fluid Tau and β-Amyloid Levels in Diabetic and Prediabetic Patients: A Meta-Analysis. Frontiers in aging neuroscience 2018, 10:271.

60. Zhang L, Chopp M, Jiang Q, Zhang Z: Role of the glymphatic system in ageing and diabetes mellitus impaired cognitive function. Stroke and vascular neurology 2019, 4(2):90–92.

61. van Dijk EJ, Breteler MM, Schmidt R, Berger K, Nilsson LG, Oudkerk M, Pajak A, Sans S, de Ridder M, Dufouil C et al: The association between blood pressure, hypertension, and cerebral white matter lesions: cardiovascular determinants of dementia study. Hypertension 2004, 44(5):625–630.

62. Dhar R, Chen Y, Hamzehloo A, Kumar A, Heitsch L, He J, Chen L, Slowik A, Strbian D, Lee JM: Reduction in Cerebrospinal Fluid Volume as an Early Quantitative Biomarker of Cerebral Edema After Ischemic Stroke. Stroke 2020, 51(2):462–467.

63. Wang YJ, Sun YR, Pei YH, Ma HW, Mu YK, Qin LH, Yan JH: The lymphatic drainage systems in the brain: a novel target for ischemic stroke? Neural Regen Res 2023, 18(3):485–491.

64. Reiber H: Cerebrospinal fluid--physiology, analysis and interpretation of protein patterns for diagnosis of neurological diseases. Multiple sclerosis (Houndmills, Basingstoke, England) 1998, 4(3):99–107.

65. Simon MJ, Iliff JJ: Regulation of cerebrospinal fluid (CSF) flow in neurodegenerative, neurovascular and neuroinflammatory disease. Biochimica et biophysica acta 2016, 1862(3):442–451.

