## Supplementary Fig. for "Genetic and Phenotypic Architecture of Brain Glymphatic System"

**Supplementary Contents List**

**Supplementary Methods.**

**Supplementary Figure 1.** Diffusion tensor image (DTI) analysis along the perivascular space (DTI-ALPS) workflow.

**Supplementary Figure 2.** Regional association plots for 14 genome-wide significant loci in the ALPS-Index GWAS.

**Supplementary Figure 3.** QQ plot for the ALPS-Index GWAS.

**Supplementary Figure 4.** Correlation analysis of ALPS Index and predictive capability of PRS Scores.

**References.**

**eMethods**

**ALPS Index calculation**

The ALPS index was calculated using a shared bash script (<https://github.com/gbarisano/alps>). Diffusion tensor metrics along the x, y, and z axes were extracted, labeled as Dxx, Dyy, and Dzz, respectively. Fractional anisotropy (FA) and diffusion rate maps (Dxx, Dyy, Dzz) were generated from the pre-processed DTI data using the FSL command-line tool "dtifit." Individual FA maps were co-registered to the JHU-ICBM FA template via linear and non-linear registration using FSL's flirt and fnirt tools. The acquired transformation matrix was applied to all the diffusivity maps (Dxx, Dyy, Dzz). Four 5 mm diameter spherical regions of interest (ROIs) were automatically placed in the areas of projection and association fibers at the level of the lateral ventricle body. For each participant, the diffusivity within the ROI template were calculated for the projection and association fibers along the x-axis (Dxx), y-axis (Dyy), and z-axis (Dzz), denoted as Dxproj, Dyproj, Dzproj, Dxassoc, Dyassoc, and Dzassoc, respectively. ALPS index was calculated as follows: $ALPS Index= \frac{mean(\mathrm{Projection}_{x},\mathrm{Association}_{x})}{mean(\mathrm{Projection}_{y},\mathrm{Association}_{z})}$. We calculated the ALPS index for the left hemisphere, the right hemisphere and the whole brain.

**Genome-Wide Association**

GWAS was conducted on the mean ALPS index for both left and right hemispheres, using linear regression to assess associations with 7,604,629 SNPs (details below). The genetic analysis was performed using the Regenie software package (version 3.4.1; <https://rgcgithub.github.io/regenie>)[1]. In Step 1, Regenie employed a nested ridge regression model to process the genetic data, dividing the genome into small blocks of 1000 SNPs each. Ridge regression was applied to each block, generating a set of local predictions, which were then combined using a second ridge regression layer to produce a single prediction. To avoid proximal contamination, chromosome-wide leave-one-chromosome-out (LOCO) processing was applied. In Step 2, the LOCO predictions from Step 1 were used as covariates to test for associations between genetic variants and the ALPS index.

For Step 1, quality-controlled imputed genotype data provided by the Wellcome Trust Centre for Human Genetics (WTCHG) was used, consisting of 7,604,629 variants covering autosomal regions with MAF >1%, missingness <10%, Hardy-Weinberg equilibrium (HWE) p > 1 × 10⁻¹⁵ (with mid-P correction), and imputation quality (mach-r²) >0.8. The data covered all 487,409 UKBB participants and was quality controlled using the plink GWAS software (version 2.0.0). To enhance computational efficiency, Regenie divided the genome into 1000-marker non-overlapping blocks (using the “--bsize” option) and used a linear mixed model (LMM) to predict and account for known confounding factors, allowing for clear assessment of the effects of individual genetic loci in Step 2. In Step 2, genetic association testing was performed using the model predictions from Step 1, alongside the same imputed genotype data and the same quality control criteria: MAF >1%, missingness <10%, HWE p > 1 × 10⁻¹⁵, and mach-r² >0.8.

Covariate adjustments were made for age (“age at assessment center,” data field 21003), sex, age × sex, age^2^, age^2^ × sex, genotype array (Axiom vs. BiLEVE), scanner site (data field 54), total intracranial volume (data field 26521), and the top 10 genetic principal components (“genetic principal components,” data field 22009) to control for population structure. Prior to running Regenie, the GWAS phenotypes were transformed using rank-based inverse normal transformation to ensure a normal distribution of the GWAS traits. Genetic inflation and heritability of the GWAS results for the ALPS index were estimated using LDSC[2].

To calculate the LD structure, identify genomic risk loci, candidate SNVs, lead SNVs, and independent significant SNVs for ALPS index, and perform variant annotation, we used the SNV2GENE pipeline in FUMA software (version 1.5.2)[3]. Independent significant SNVs were defined as those meeting genome-wide significance thresholds (nominal p-value threshold of 5 × 10^-8^, significance threshold of p < 0.05) and were independent of each other at an r² < 0.6 level. Lead SNVs were defined more strictly as independent SNVs at r² < 0.1. Genomic risk loci were defined by FUMA as regions containing physically proximal or overlapping independent signals, as well as lead SNVs. First, independent signals SNVs dependent on each other at r² < 0.1 were grouped into the same genomic risk locus. Next, independent significant SNPs located within 250 kb of each other were merged into a single genomic risk locus. The reference panel used was the UKBB release2b 10k White British cohort. Variants were annotated using ANNOVAR[4] software (version 2017-07-17), embedded within FUMA. FUMA also cross-referenced candidate SNVs identified from our GWAS summary statistics with the GWAS catalog (version e0_r2022-11-29). Functional annotations of variants included predicted pathogenicity (CADD score) and regulatory effects (RegulomeDB score and chromatin state).

**Conditional and Joint Association Analysis**

Using Conditional and joint multiple-SNP analysis using Genome-wide Complex Trait Analysis (GCTA)-COJO, conditional and joint analyses were conducted to identify independent signals within significant loci. GWAS summary statistics and LD structure from the UK Biobank reference panel were used, applying a MAF threshold of 1% and a 10 Mb LD window. Independent SNPs were identified with an r² < 0.6, and lead SNPs with r² < 0.1.

To investigate whether a genome-wide significant locus consists of multiple independent signals, we performed Conditional and joint multiple-SNP analysis using Genome-wide Complex Trait Analysis (GCTA)-COJO[5]. The COJO analysis utilized GWAS summary statistics and the linkage disequilibrium (LD) structure from a reference panel to iteratively condition on the top SNPs in the GWAS summary data. Quality-controlled imputed genotype data from the Wellcome Trust Centre for Human Genetics (WTCHG) in the UK Biobank was used as the reference panel for LD estimation. We set a MAF threshold of 0.01 and applied a 10 Mb LD window. Independent significant SNPs were identified using a stepwise selection model (with the --cojo-slct option), while setting the collinearity threshold to 0.2. The conditional joint analysis was conducted across chromosomes 1 to 22.

**Candidate Gene Identification and Annotation**

We employed a positional mapping approach to link genome-wide significant loci to genes using the default settings in FUMA[3]. Specifically, variants were mapped within a 10 kB window surrounding genes coding for known proteins in the human reference genome assembly (GRCh37/hg19).

For gene-based association analysis, we utilized the default parameters of Multi-marker Analysis of GenoMic Annotation (MAGMA)[6] within FUMA to aggregate association signals from the SNP level to the gene level. A total of 19,141 protein-coding genes were analyzed in MAGMA. The mean chi-square (χ²) of the GWAS summary statistics was used to calculate gene-based p-values. The significance threshold for the p-values was determined using the Bonferroni correction method, with a threshold of 2.61 × 10⁻⁶, obtained by dividing 0.05 by the total number of genes (19,141).

**Functional Annotation of Susceptible Genes**

We further sought to identify candidate genes influencing ALPS Index phenotypic variation using an integrative approach supported by multiple lines of evidence. Genes annotated by more than three methods were classified as candidate genes for ALPS Index. To investigate the functional characteristics of these candidate genes, we performed gene enrichment analysis using the DAVID platform (https://david.ncifcrf.gov/)[7], which integrates multiple bioinformatics resources through annotations to identify gene sets significantly enriched in various biologically relevant categories. Three Gene Ontology (GO) categories—Molecular Function, Cellular Component, and Biological Process—were analyzed, and the FAT categories in DAVID were employed to refine the enrichment results by filtering out broad, non-informative GO terms. This approach allowed us to focus on more specific, relevant categories, thereby providing a clearer understanding of the biological roles of the identified genes. The significance threshold was set at a false discovery rate (FDR) of < 0.05, using the Benjamini-Hochberg correction for multiple testing.

**Cell-Specific Susceptibility Analysis**

To identify cellular targets associated with the ALPS index, we applied a cell-specific susceptibility scoring approach using the scDRS package (version 1.0.3). The scDRS algorithm, developed by Zhang et al., is a novel method that links single-cell RNA sequencing data to polygenic disease risk at single-cell resolution[8]. The process comprises the following steps: (i) ALPS-Index Gene Set Construction: Using GWAS summary statistics related to the ALPS index, scDRS identifies a set of putative ALPS Index-associated genes. (ii) Score Calculation: For each cell in the single-cell RNA sequencing dataset, scDRS calculates a raw score based on the expression levels of the ALPS Index -related genes. Additionally, scDRS generates a Monte Carlo sample of B raw control scores for each cell (with a default B = 1000). (iii) Normalization and P-Value Calculation: Normalization is performed at both the gene set and cell levels. Following normalization, scDRS calculates an association p-value by comparing the empirical distribution of normalized scores for each cell to the normalized control scores. These p-values can also be used for data visualization and further statistical inference.

**Disease Association Analysis**

To investigate the associations between candidate genes and known diseases, we utilized the Enrichr tool[9] (https://maayanlab.cloud/Enrichr/) and selected the GWAS Catalog 2023 database for analysis. The GWAS Catalog 2023 (https://www.ebi.ac.uk/gwas/) compiles results from numerous GWAS related to various diseases, allowing us to evaluate the genetic associations of candidate genes across a range of conditions. The significance of enrichment was assessed by calculating enrichment scores and adjusted p-values using the Benjamini-Hochberg correction. The significance threshold was set at a FDR of < 0.05.

**Enrichment of Drug Target Genes**

We collected all the druggable genes mapped to Entrez gene IDs using the DGIdb database[10]. In addition, we referred to Finan et al. who linked GWAS loci of complex diseases to druggable genes[11]. From DGIdb, we extracted 5012 potential drug target genes (eTable 1 in Supplement 2) and identified 4463 druggable genes (eTable 2 in Supplement 2) from the study of Finan et al. To ensure the reliability of the selected genes and their potential as valid drug targets, we further screened 2,587 genes that were validated by two sources and had official names assigned by the Human Genome Nomenclature Committee (HGNC).

Within this screened gene set, any gene identified by three or more methods together was identified as a druggable gene associated with ALPS Index. Subsequently, we searched the DrugBank (https://go.drugbank.com) and ClinicalTrials (https://www.clinicaltrials.gov) databases to assess the drug development status and clinical development activities of these identified drug target genes[12].

**Association Analysis Between ALPS Index and Five Major Categories of Non-Imaging Phenotypes**

In this study, we first used the ALPS index, rank-transformed using inverse normal transformation from 36,997 participants, to perform a phenotype association analysis. This analysis aimed to investigate the associations between the ALPS index and all available risk factors, evaluating both effect size and statistical significance. We analyzed 2,121 phenotypes obtained from the UKBB, covering five groups of variable types: (i) Biomarkers and Physical Measurements, (ii) Medical Conditions and Treatments, (iii) Cognitive and Mental Health, (iv) Lifestyle and Social Factors, and (v) Sex-Specific Factors. These variables were derived from five categories in the UK Biobank dataset: Additional Exposures, Assessment Centre, Biological Samples, Health-Related Outcomes, and Online Follow-Up.

The PheWAS associations were tested using the PHESANT package in R. The automated rule-based approach employed by PHESANT has been described in detail in prior publications[13]. This step, referred to as “ALPS Index-PheWAS,” was based on decision rules that classified each variable into one of four data types: continuous, ordered categorical, unordered categorical, or binary.

In the “ALPS Index-Association Analysis” step, the rank-transformed ALPS index was set as the independent variable, and the selected factors were treated as dependent variables, with age, sex, and assessment center included as covariates in the model. To enable direct comparison between results from linear and logistic regression models, standardized regression coefficients were estimated as effect sizes (β), and for binary outcome variables, log-transformed odds ratios (OR) were reported. All analyses employed two-sided statistical tests. The results of 2,121 associations were corrected for multiple testing using the FDR correction, and results with p_FDR_ < 0.05 were summarized for subsequent validation analyses.

**Polygenic Risk Score Validation of ALPS Index Associations Across Five Phenotype Categories**

In the validation process, we first calculated the PRS for the ALPS index in participants from the UKBB who did not have a directly measured ALPS index. We used data from the UKBB’s prospective cohort study, which recruited over 502,364 UK participants between 2006 and 2010, with genotype data available for 488,127 individuals. Detailed genotyping and quality control procedures have been previously described in earlier publications. We excluded SNPs with a call rate below 95% and MAF below 0.1%. Participants of recent British ancestry were selected based on self-reported information and principal component analysis of the genotype data. Individuals with more than one inferred third-degree relative and those with a directly measured ALPS index were excluded. After quality control, 370,920 participants were included in the analysis.

PRS for the ALPS index were calculated for these 370,920 individuals using PRS-CS[14], based on the GWAS summary statistics from 36,111 participants of White British ancestry with available ALPS data. Details of the GWAS summary data generation process are provided in subsequent sections. The ALPS index-PRS was set as the independent variable, and the factors identified in the “ALPS Index-Association Analysis” step (p_FDR_ < 0.05) were set as the dependent variables. Age, sex, genotyping array, the top 10 genetic principal components, and assessment center were included as covariates in the model for phenotype association analysis. This step, referred to as “ALPS Index-PRS-PheWAS,” was conducted using the PHESANT package in R. To enable direct comparison between linear and logistic regression model results, standardized regression coefficients were used as effect sizes (β), and for binary outcome variables, log-transformed OR were reported. Two-sided statistical tests were employed for all analyses. The 181 association results were corrected using the FDR correction.

**Validation with Neurological Diseases Using Mendelian Randomization (MR) and Cox**

To further validate the significant negative associations identified in the association analysis, we used multivariable Cox proportional hazards regression models to investigate the relationship between the ALPS Index and the incidence of common neurological diseases, including AD, MS, cerebral infarction, and stroke (unspecified as hemorrhage or infarction). Follow-up time was calculated in person-years, beginning from the second recruitment visit (instance 2) until the first diagnosis of a neurological disease, death, loss to follow-up, or the date of the last recorded hospital admission, whichever occurred first. The model was adjusted for age, sex, Townsend deprivation index, and BMI. The proportional hazards assumption was evaluated using the Schoenfeld residual test. In the primary analysis, participants diagnosed with neurological diseases prior to baseline were excluded.

Following this, MR[15] was employed to assess potential causal relationships between the ALPS Index and common neurological diseases, including AD, MS, and stroke (eTable 20 in Supplement 2). Within a two-sample MR framework, we explored risk factors associated with ALPS index. MR leverages genetic variants as instrumental variables to infer causal relationships between exposures and outcomes. Genetic instruments were selected from GWAS summary statistics using a p-value threshold of 5E-8. These SNPs were then clumped based on a 10,000 kb distance and an LD r² threshold of 0.001 to obtain independent instruments. Only SNPs present in both the phenotype and PD GWAS datasets were retained. The primary MR method used was inverse variance weighting (IVW), which assumes no measurement error for the association between genetic variants and exposure, thus avoiding bias in causal effect estimation. Potential weak instrument bias, indicated by F-statistics, was addressed by selecting instruments with F-statistics greater than 10 for subsequent analyses. When causal effects were detected using the IVW method. These robust approaches account for violations of MR assumptions under different conditions, enabling effective causal effect estimation. In addition to these robust MR methods, sensitivity tests, including Cochran’s Q statistic, were conducted to detect heterogeneity in instrument-specific causal estimates and to assess horizontal pleiotropy.

**Data and code availability**

Data: The data supporting the findings of this study are available in the following resources. Data from UK Biobank DWI scans were collected according to the published protocol (https://biobank.ctsu.ox.ac.uk/crystal/refer.cgi?id=2367). Permission to use the UK Biobank Resource was obtained via a material transfer agreement as part of Data Access Application 221671. All imaging data, phenotypes, and genetic data are made available by UK Biobank via their standard data access procedure (http://www.ukbiobank.ac.uk/register-apply). The MRI data processing pipelines used in the UK Biobank project can be found at http://www.fmrib.ox.ac.uk/ukbiobank or on GitLab at https://git.fmrib.ox.ac.uk/falmagro/UK_biobank_pipeline_v_1.

Code: The following software packages were used in this work:

- FMRIB Software Library (FSL) v6.0.7.11 (https://fsl.fmrib.ox.ac.uk/fsl/fslwiki) was used for image analysis and statistical tools specifically designed for brain imaging data.
- mrtrix3 v3.0.4 (https://www.mrtrix.org/) provided tools for advanced diffusion MRI processing, allowing for tractography and fiber orientation estimation.
- ALPS index (https://github.com/gbarisano/alps), calculated using a shared bash script, was applied to assess cerebrospinal fluid flow dynamics.
- Regenie v3.4.1 (https://rgcgithub.github.io/regenie) was used for genome-wide association studies (GWAS) to handle large-scale genetic data and estimate polygenic risk scores.
- FUMA v1.5.2 (https://fuma.ctglab.nl) served as a post-GWAS annotation tool, enabling functional mapping and annotation of genetic associations.
- GCTA-COJO v1.94.1 (https://cnsgenomics.com/software/gcta) was used for conditional and joint association analyses to estimate genetic heritability.
- MAGMA v1.08 (https://ctg.cncr.nl/software/magma) enabled gene-set analysis to identify relevant gene pathways associated with phenotypes.
- GWAS Catalog v2022-11-29 (https://www.ebi.ac.uk/gwas) provided a comprehensive database of published genome-wide association studies to compare and validate findings.
- ANNOVAR v2017-07-17 (https://annovar.openbioinformatics.org) facilitated variant annotation and functional annotation of genetic variants.
- FUSION (http://gusevlab.org/projects/fusion) was utilized for transcriptome-wide association studies, linking gene expression with traits.
- COLOC v5.1.0 (https://cran.r-project.org/web/packages/coloc) allowed colocalization analysis to identify shared genetic signals between traits.
- DAVID v2023q4 (https://david.ncifcrf.gov) provided functional annotation for gene lists, aiding in biological interpretation of gene sets.
- scDRS v1.0.3 (https://github.com/martinjzhang/scDRS) was used for single-cell disease relevance scoring to explore cell-type specific relevance of genetic associations.
- Enrichr v2023-6-8 (https://maayanlab.cloud/Enrichr) offered gene list enrichment analysis to identify biological pathways and functions.
- GWAS Catalog 2023 (https://www.ebi.ac.uk/gwas) offered an updated repository of GWAS studies for identifying previously associated genetic variants.
- DGIdb v5.0.7 (https://www.dgidb.org) provided drug-gene interaction data, useful for identifying potential therapeutic targets.
- DrugBank (https://go.drugbank.com), accessed in August 2024, was consulted for drug information and interactions.
- ClinicalTrials (https://www.clinicaltrials.gov), accessed in August 2024, provided data on ongoing clinical trials related to identified targets.
- PHESANT v1.1 (https://github.com/MRCIEU/PHESANT) was used for phenome-wide association studies (PheWAS) to explore associations between genetic variants and phenotypes.
- PRS-CS v1.1.0 (https://github.com/getian107/PRScs) generated polygenic risk scores using a Bayesian approach to improve prediction accuracy.
- LDSC v1.0.1 (https://github.com/bulik/ldsc) was applied for linkage disequilibrium score regression, estimating heritability and genetic correlation.
- MR v0.5.6 (https://github.com/mrcieu/TwoSampleMR) was used for Mendelian randomization analyses to assess causal relationships between traits.

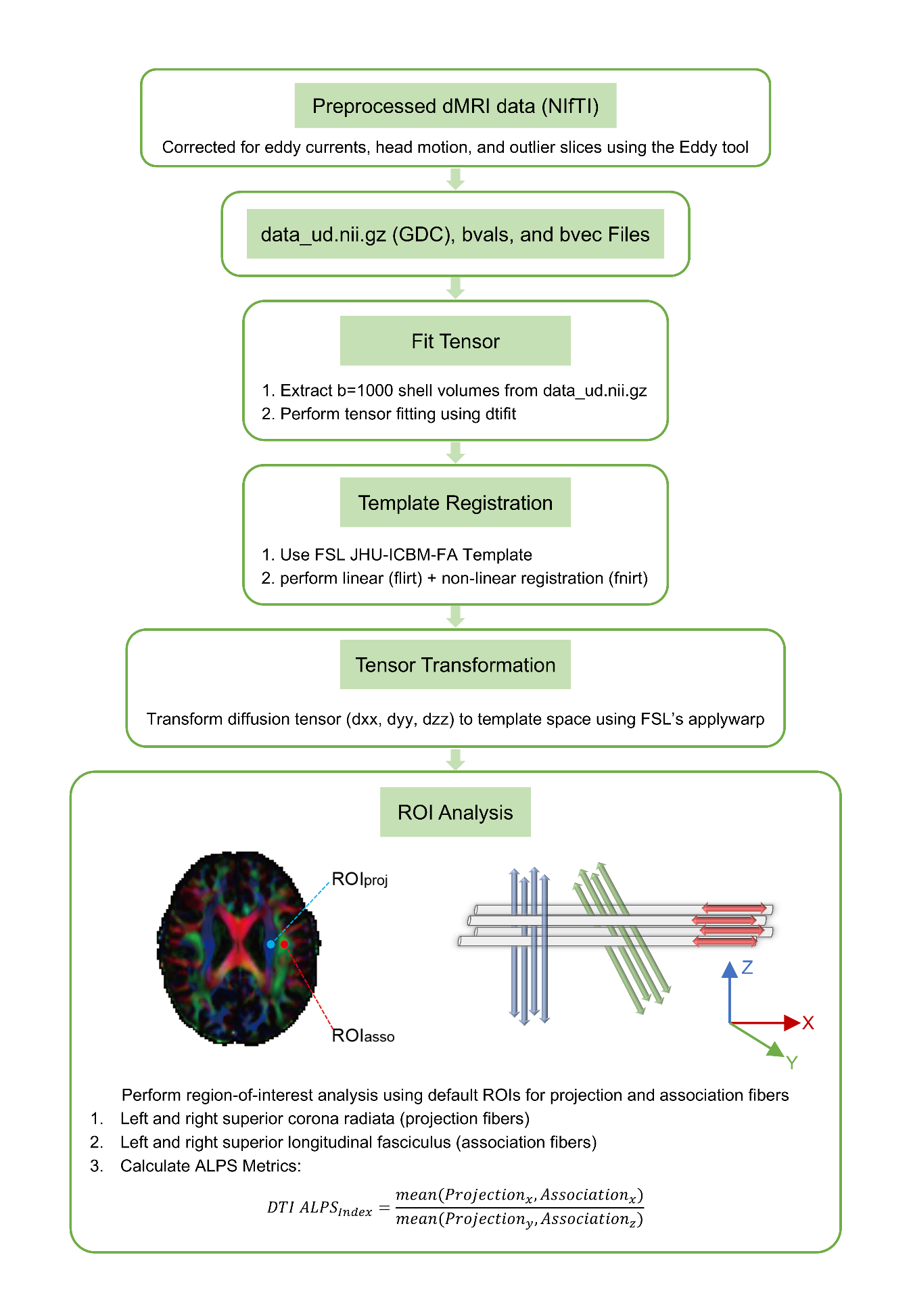
**Supplementary Figure 1.** Diffusion tensor image (DTI) analysis along the perivascular space (DTI-ALPS) workflow.

dtifit: Command provided by FSL used to perform tensor fitting and extract diffusivity measures; flirt: Command provided by FSL for linear image registration, used to align individual images to a standardized template space; fnirt: Non-linear registration tool from FSL used to further refine the alignment to the FSL JHU-ICBM-FA template; Dxx indicates diffusivity along the perivascular space, signifying left to right directional diffusion; Dyy, diffusivity along projection fibers, indicating superior to inferior directional diffusion; Dzz, diffusivity along association fibers, depicting anterior to posterior directional diffusion; FSL, Oxford Centre for Functional Magnetic Resonance Imaging of the Brain Software Library; and ROI, region of interest.

**Supplementary Figure 2.** Regional association plots for 14 genome-wide significant loci in the ALPS-Index GWAS.

| 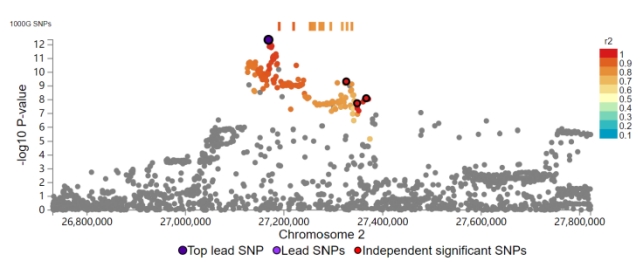 | 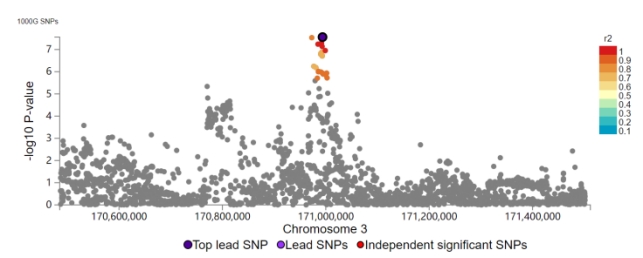 |
| --- | --- |
| 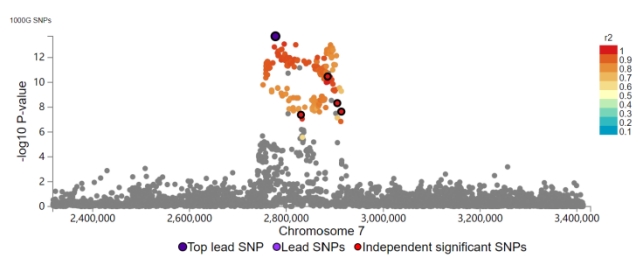 | 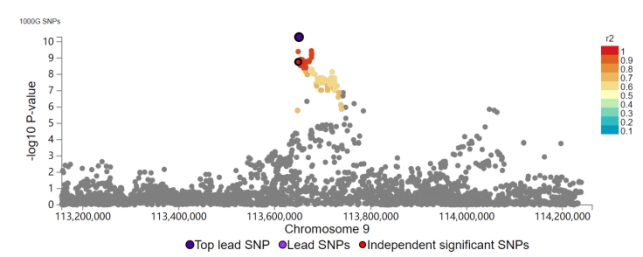 |
| 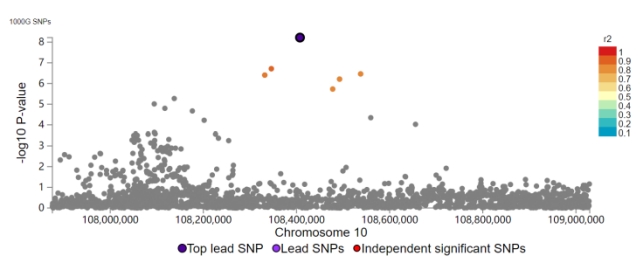 | 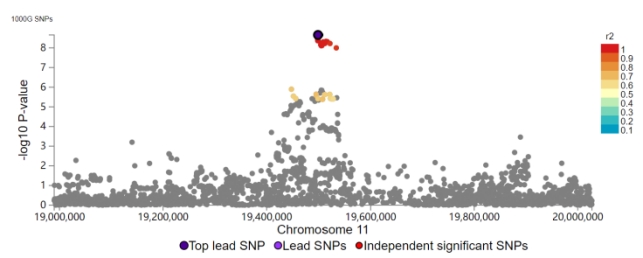 |
| 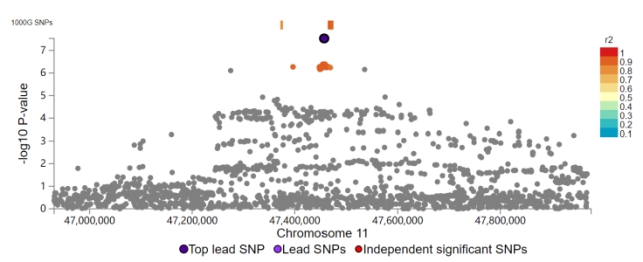 | 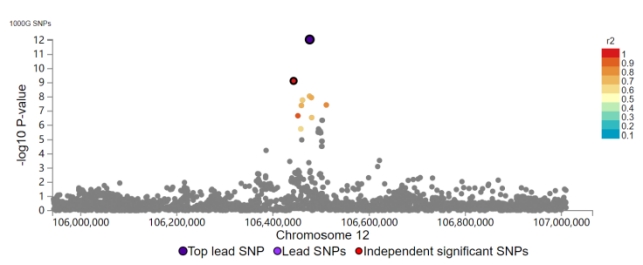 |
| 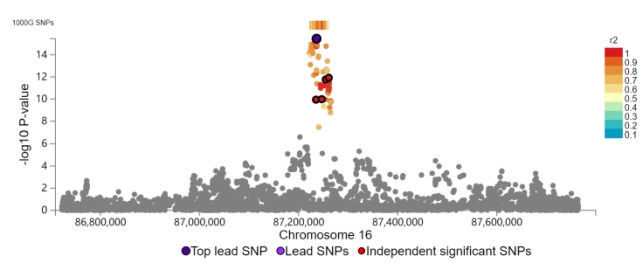 | 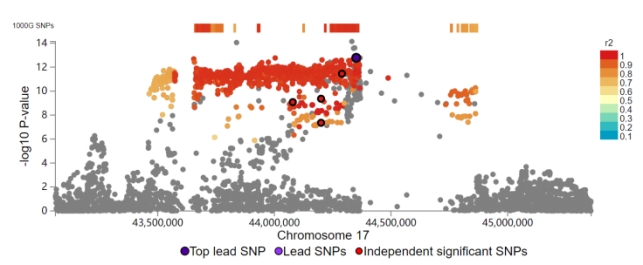 |
| 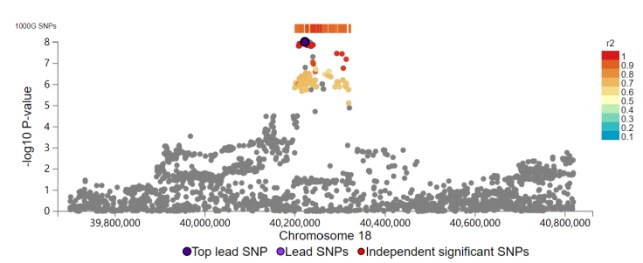 | 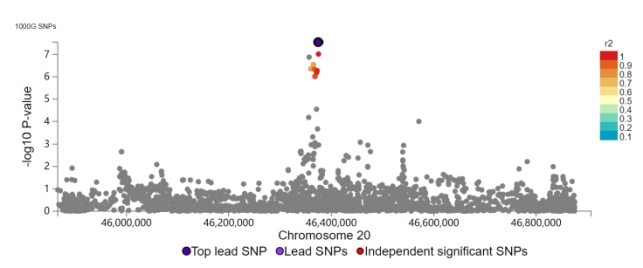 |
| 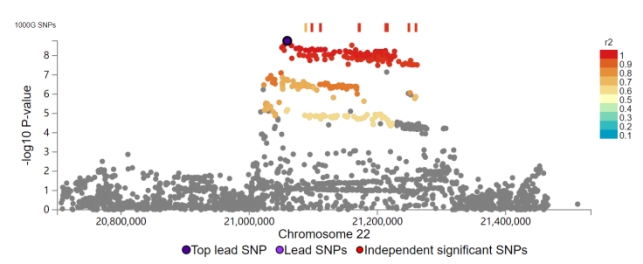 | 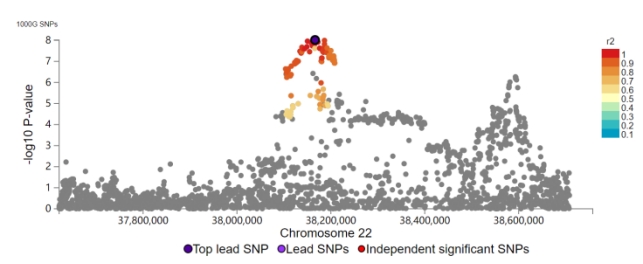 |

**Supplementary Figure 3.** QQ plot for the ALPS-Index GWAS.

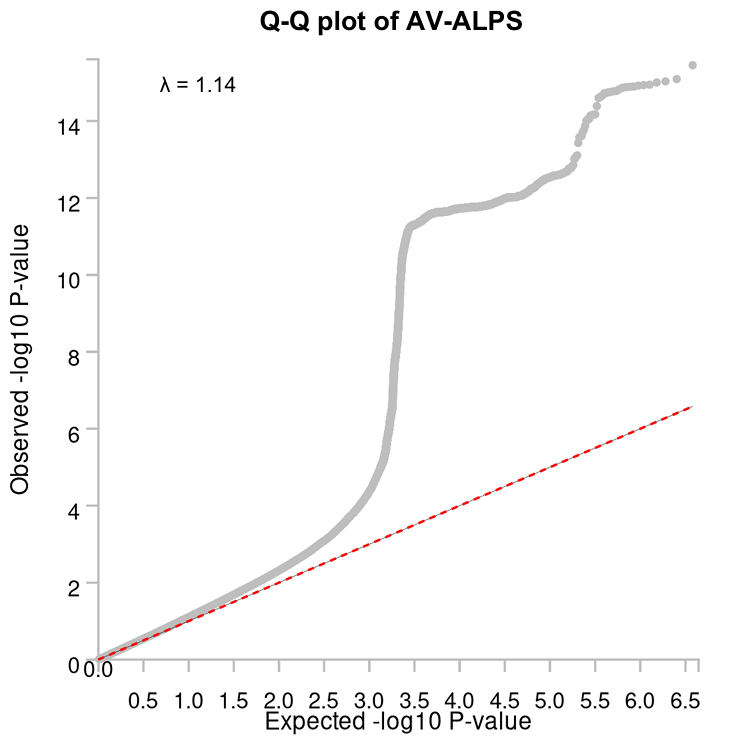

This quantile-quantile (Q-Q) plot illustrates the distribution of observed versus expected -log_10_ P-value from the GWAS for the average ALPS index (AV-ALPS). λ: The genomic inflation factor.

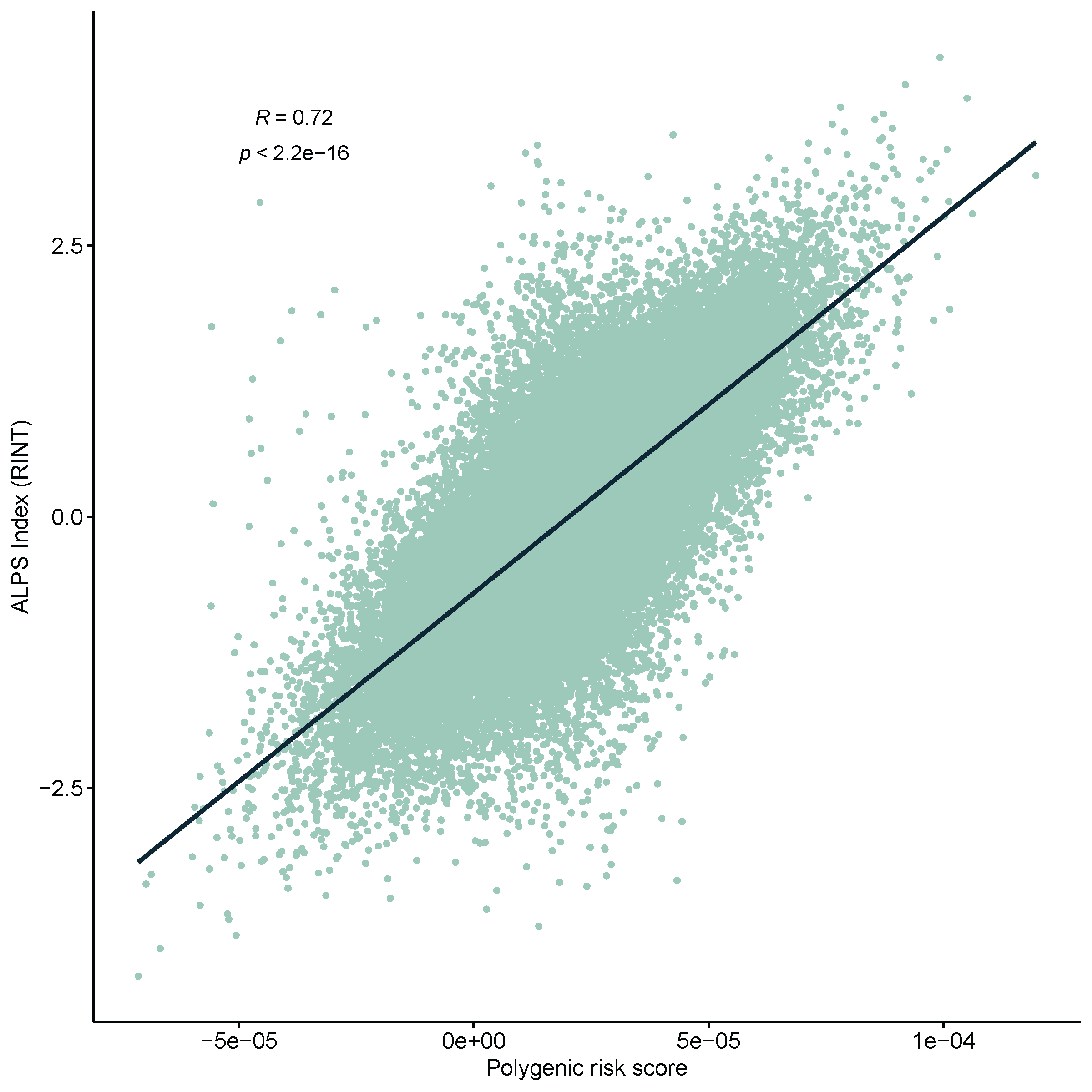
**Supplementary Figure 4.** Correlation analysis of ALPS Index and predictive capability of PRS Scores.

The scatter plot shows the correlation between the polygenic risk score (PRS) and the rank-inverse normal transformed (RINT) ALPS Index. The Pearson correlation coefficient *R* = 0.72 with p < 2.2 × 10^−16^ suggests a strong positive correlation, indicating that higher PRS is associated with higher ALPS Index values.

**References**

1.	Mbatchou J, Barnard L, Backman J, Marcketta A, Kosmicki JA, Ziyatdinov A, Benner C, O'Dushlaine C, Barber M, Boutkov B *et al*: **Computationally efficient whole-genome regression for quantitative and binary traits**. *Nature genetics* 2021, **53**(7):1097-1103.

2.	Bulik-Sullivan BK, Loh PR, Finucane HK, Ripke S, Yang J, Patterson N, Daly MJ, Price AL, Neale BM: **LD Score regression distinguishes confounding from polygenicity in genome-wide association studies**. *Nature genetics* 2015, **47**(3):291-295.

3.	Watanabe K, Taskesen E, van Bochoven A, Posthuma D: **Functional mapping and annotation of genetic associations with FUMA**. *Nature communications* 2017, **8**(1):1826.

4.	Wang K, Li M, Hakonarson H: **ANNOVAR: functional annotation of genetic variants from high-throughput sequencing data**. *Nucleic acids research* 2010, **38**(16):e164.

5.	Yang J, Ferreira T, Morris AP, Medland SE, Madden PA, Heath AC, Martin NG, Montgomery GW, Weedon MN, Loos RJ *et al*: **Conditional and joint multiple-SNP analysis of GWAS summary statistics identifies additional variants influencing complex traits**. *Nature genetics* 2012, **44**(4):369-375, s361-363.

6.	de Leeuw CA, Mooij JM, Heskes T, Posthuma D: **MAGMA: generalized gene-set analysis of GWAS data**. *PLoS computational biology* 2015, **11**(4):e1004219.

7.	Sherman BT, Hao M, Qiu J, Jiao X, Baseler MW, Lane HC, Imamichi T, Chang W: **DAVID: a web server for functional enrichment analysis and functional annotation of gene lists (2021 update)**. *Nucleic acids research* 2022, **50**(W1):W216-w221.

8.	Zhang MJ, Hou K, Dey KK, Sakaue S, Jagadeesh KA, Weinand K, Taychameekiatchai A, Rao P, Pisco AO, Zou J *et al*: **Polygenic enrichment distinguishes disease associations of individual cells in single-cell RNA-seq data**. *Nature genetics* 2022, **54**(10):1572-1580.

9.	Xie Z, Bailey A, Kuleshov MV, Clarke DJB, Evangelista JE, Jenkins SL, Lachmann A, Wojciechowicz ML, Kropiwnicki E, Jagodnik KM *et al*: **Gene Set Knowledge Discovery with Enrichr**. *Current protocols* 2021, **1**(3):e90.

10.	Cannon M, Stevenson J, Stahl K, Basu R, Coffman A, Kiwala S, McMichael JF, Kuzma K, Morrissey D, Cotto K *et al*: **DGIdb 5.0: rebuilding the drug-gene interaction database for precision medicine and drug discovery platforms**. *Nucleic acids research* 2024, **52**(D1):D1227-d1235.

11.	Finan C, Gaulton A, Kruger FA, Lumbers RT, Shah T, Engmann J, Galver L, Kelley R, Karlsson A, Santos R *et al*: **The druggable genome and support for target identification and validation in drug development**. *Science translational medicine* 2017, **9**(383).

12.	Knox C, Wilson M, Klinger CM, Franklin M, Oler E, Wilson A, Pon A, Cox J, Chin NEL, Strawbridge SA *et al*: **DrugBank 6.0: the DrugBank Knowledgebase for 2024**. *Nucleic acids research* 2024, **52**(D1):D1265-d1275.

13.	Millard LAC, Davies NM, Gaunt TR, Davey Smith G, Tilling K: **Software Application Profile: PHESANT: a tool for performing automated phenome scans in UK Biobank**. *International journal of epidemiology* 2018, **47**(1):29-35.

14.	Ge T, Chen CY, Ni Y, Feng YA, Smoller JW: **Polygenic prediction via Bayesian regression and continuous shrinkage priors**. *Nature communications* 2019, **10**(1):1776.

15.	Hemani G, Zheng J, Elsworth B, Wade KH, Haberland V, Baird D, Laurin C, Burgess S, Bowden J, Langdon R *et al*: **The MR-Base platform supports systematic causal inference across the human phenome**. *eLife* 2018, **7**.
